# Altered sex differences in hippocampal subfield volumes in schizophrenia

**DOI:** 10.1101/2023.01.26.23284945

**Authors:** Claudia Barth, Stener Nerland, Kjetil N. Jørgensen, Beathe Haatveit, Laura A. Wortinger, Ingrid Melle, Unn K. Haukvik, Torill Ueland, Ole A. Andreassen, Ingrid Agartz

## Abstract

**Objective:** The hippocampus is a heterogenous brain structure that differs between the sexes and has been implicated in the pathophysiology of psychiatric illnesses. Here, we explored sex and diagnostic group differences in hippocampal subfield volumes, in individuals with schizophrenia spectrum disorder (SZ), bipolar disorders (BD) and healthy controls.

**Methods:** 1,521 participants underwent T1-weighted magnetic resonance imaging (SZ, n = 452, mean age 30.7 ± 9.2 [SD] years, males 59.1%; BD, n = 316, 33.7 ± 11.4, 41.5%; healthy controls, n = 753, 34.1 ± 9.1, 55.6%). Total hippocampal, subfield, and intracranial volumes were estimated with Freesurfer (v6.0.0). Analysis of covariance and multiple regression models were fitted to examine sex-by-diagnostic (sub)group interactions in volume. In SZ and BD, separately, associations between volumes and clinical as well as cognitive measures were examined between the sexes using regression models.

**Results:** Significant sex-by-group interactions were found for the total hippocampus, dentate gyrus, molecular layer, presubiculum, fimbria, HATA, and CA4, indicating a larger volumetric deficit in male patients relative to female patients when compared with same-sex healthy controls. Subgroup analyses revealed that this interaction was driven by males with schizophrenia. Effect sizes were overall small (partial η^2^ < 0.02). We found no significant sex differences in the associations between hippocampal volumes and clinical or cognitive measures in SZ and BD.

**Conclusions:** Using a well-powered sample, our findings indicate that the pattern of morphological sex differences in hippocampal subfields is altered in individuals with schizophrenia relative to healthy controls, due to higher volumetric deficits in males.

## Introduction

Schizophrenia is a severe, multifactorial brain disorder with prominent sex differences in prevalence and presentation ^1^. Relative to females, males have a 40% higher likelihood of developing schizophrenia and tend to suffer from more severe forms of the illness, including worse long-term outcomes and more pronounced and persistent negative symptoms ^1^. Sex differences in the prevalence and presentation of bipolar disorders have also been reported, but a clear consensus is lacking ^2^. Despite reported sex differences in disease manifestation, it remains unknown whether and how these sex differences are reflected in the brain. Acquiring such knowledge would constitute a critical step toward mechanistic models explaining sex differences in disease susceptibility.

One brain structure that seems to differ between the sexes and has been implicated in the pathophysiology of both schizophrenia and bipolar disorders is the hippocampus ^3^. Located in the medial temporal lobe, the hippocampus is a geminate limbic brain structure with a critical role in learning ^4^, memory formation ^5^, and mood-regulation ^6^. These hippocampal-dependent functions differ between the sexes ^7^ and are affected in schizophrenia and bipolar disorders. For instance, using functional magnetic resonance imaging (MRI), lower hippocampal activation during encoding and recognition tasks has been associated with alterations in declarative memory performance in schizophrenia ^8, 9^. Similarly, structural MRI studies have shown smaller hippocampal volumes in both schizophrenia and bipolar disorders ^10, 11^, with greater volumetric deficits in schizophrenia ^12^. Hippocampal volume alterations have also been associated with cognitive impairment ^13, 14^, and thus appear functionally relevant ^15^. However, sex differences in these associations have scarcely been studied, as sex is frequently used as a covariate and not as a discovery variable ^16, 17^.

The hippocampus is, however, not a uniform structure; rather it consists of heterogeneous subfields which serve distinct functions. Using a combination of ultra-high-resolution *ex vivo* MRI data and conventional *in vivo* MRI scans, Iglesias and colleagues created a probabilistic labeling algorithm. This method segments twelve hippocampal subfields, namely the parasubiculum, presubiculum, subiculum, cornu ammonis (CA)1, CA3, CA4, granule cells in the molecular layer of the dentate gyrus (GC-ML-DG), hippocampal-amygdaloid transition area (HATA), fimbria, molecular layer, hippocampal fissure and hippocampal tail ^18^. Functional separations of hippocampal subfields have been proposed, particularly in the context of declarative memory. For instance, DG and CA3 are thought to play a crucial role in pattern separation, the flexible distinction between multiple, highly similar memories ^19^. Similarly, mechanisms in CA3, CA1 and subiculum may be critical for pattern completion, the retrieval of full memories from incomplete input. Deficits in these subfields have been proposed to results in spurious or false memory associations, creating a susceptibility to psychosis ^19^.

Meta-analyses in schizophrenia and bipolar disorders suggest lower volumes in all hippocampal subfields, predominantly in schizophrenia, with the largest effects in the CA1, CA3, CA4, GC-ML-DG and subiculum ^20^. A recent large-scale collaborative study in bipolar disorder found similarly smaller volumes in nine of 12 hippocampal subfields, excluding the fimbria, fissure and parasubiculum, in individuals with bipolar disorders relative to healthy controls ^21^. Directly comparing both disorders, previous studies found that the magnitudes of lower hippocampal subfield volumes were again greater in schizophrenia than in bipolar disorder ^22^. According to findings by Haukvik and colleagues, the left CA2 and right subiculum were smaller in schizophrenia than bipolar disorder, suggesting differential patterns of volumetric deficits depending on diagnosis ^20^.

Although diagnostic group differences in hippocampal subfield volume have been extensively demonstrated with varying degree of specificity, sex differences in hippocampal volumes are still debated. A recent study in over 1,500 healthy adults revealed region-specific sex differences in hippocampal subfield volumes, with larger volumes in males relative to females, most prominently in the fimbria and parasubiculum ^23^. Similar studies in schizophrenia and bipolar disorders are currently lacking. While emerging evidence suggests that lower hippocampal volumes are only present in males and absent in females with schizophrenia ^24, 25^, subfield-specific sex differences in schizophrenia are yet to be reported. Investigating the effect of sex on structural and functional hippocampal subfield alterations in schizophrenia and bipolar disorders may advance our understanding of the pathophysiology of these disorders and their relation to sex.

Here, we aimed to explore sex differences in hippocampal subfield volumes in a large sample of individuals with schizophrenia spectrum disorders and bipolar disorders and healthy controls. We further examined the associations between symptom profiles, cognitive measures as well as medication intake on hippocampal volumes by sex and diagnostic groups. In the current study, we focused on four cognitive domains, that have been previously linked to hippocampal functioning ^13, 14^, may vary between the sexes ^26-28^ and may be impaired in schizophrenia and bipolar disorders ^29^, namely processing speed, working memory, verbal learning and verbal memory. Due to a lack of consistent evidence on sex differences in hippocampal total and subfield volumes in schizophrenia and bipolar disorders, the analyses were exploratory by nature.

## Methods and materials

### Participants

The sample of 768 individuals with schizophrenia spectrum disorders (SZ) and bipolar disorders (BD), and 753 healthy controls is part of the ongoing Thematically-Organized-Psychosis (TOP) study, Oslo, Norway. Individuals with SZ and BD were recruited from in- and outpatient psychiatric units covering specific catchment areas in the greater Oslo area. Healthy controls were recruited from the national population register in the same catchment area. All participants provided written informed consent. The study was approved by the Regional Committee for Research Ethics and the Norwegian Data Inspectorate, and carried out in accordance with the Declaration of Helsinki. For details on inclusion criteria, see supplementary materials Note 1.

### Clinical assessment

Clinical characterization was conducted by trained clinical psychologists, psychiatrists, and medical doctors. Clinical diagnoses were established according to the Structured Clinical Interview for Diagnostic and Statistical Manual of Mental Disorder (DSM)-IV axis 1 disorder (SCID-I), module A-E ^30^. Healthy controls were evaluated with the Primary Care Evaluation of Mental Disorders (Prime-MD)^31^, and participants who presented with a history of severe mental illness or had mental disorders in their close family were excluded. The Positive and Negative Syndrome Scale (PANSS) ^32^ was used to assess the presence and severity of psychotic symptoms, and the Inventory for Depressive Symptomatology (IDS) ^33^ and the Young Mania Rating Scale (YMRS) ^34^ to evaluate affective symptoms. General functioning in all participants was measured with the Global Assessment of Function (GAF) scale, split version ^35^.

Clinical diagnoses were: (1) *schizophrenia spectrum disorders* (SZ; n = 452): schizophrenia [SCZ, DSM-IV 295.1, 295.3, 295.6, 295.9; n = 245], schizophreniform [DSM-IV 295.4; n = 31], schizoaffective [SCZ-AF, DSM-IV 295.7; n = 59], other psychotic disorders [OPD, DSM-IV 297.1, 298.8, 298.9; n = 117], (2) *bipolar disorders* (BD, n = 316): bipolar I [DSM-IV 296.0-7; n = 188], bipolar II [DSM-IV 296.89; n = 113], bipolar not otherwise specified [DSM-IV 296.8; n = 15]. Most individuals with BD presented with a history of psychotic features (60.4 %) and were investigated in their euthymic phase (87.3 %, i.e., YMRS < 8 and IDS ≤ 13)^36^.

Age of onset for SZ was defined as age at first psychosis. For BD, age of onset was defined as the age when the first mood episode occurred. Mood episodes include manic or hypomanic episodes, mixed episodes and depressive episodes. We set the lowest age for either of these mood episodes as the age of onset. Duration of illness was calculated based on age at MRI scan minus age of onset.

### Cognitive assessment

Clinical psychologists and trained personnel administered cognitive assessments of the clinical groups and healthy controls, respectively. Between 2004-2019, the cognitive assessment was performed using two different cognitive test batteries. 729 participants were evaluated with battery 1, a standardized battery described in detail elsewhere ^26^. Starting from 2012, participants (n = 722) were assessed using a licensed translated version of the Measurement and Treatment Research to Improve Cognition in Schizophrenia (MATRICS) Consensus Cognitive Battery (MCCB) ^37^. In the current study, we focused on four cognitive domains: (1) *processing speed*, measured with Digit Symbol Coding Test from the Wechsler Adult Intelligence Scale-III (WASI-III) or Brief Assessment of Cognition in Schizophrenia (BACS); (2) *working memory*, measured with the Letter-Number Sequencing Test from WASI-III or the Letter-Number Sequencing Test from the MCCB; (3) *verbal learning and (4) verbal memory* measured with the California Verbal Learning Test (CVLT) or the Hopkins Verbal Learning Test, Revised (HVLT-R, for details see Table 1). A total of 67 participants did not have cognitive data. Current intelligence quotient (IQ) was assessed using the Wechsler Abbreviated Scale of Intelligence (WASI, four subtests).

**Table 1.** Demographics and clinical measures of the study sample.

|  | Healthy Controls |  |  | BD |  |  | SZ |  |  | Dx Differences |  |
| --- | --- | --- | --- | --- | --- | --- | --- | --- | --- | --- | --- |
|  | Female<br>N = 334 | Male<br>N = 419 | p-value | Female<br>N = 185 | Male<br>N = 131 | p-value | Female<br>N = 185 | Male<br>N = 267 | p-value | Female,<br>p-value | Male,<br>p-value |
| **Age at Scan** (years) | 33.4<br>[26.3, 40.5] | 33.1<br>[27.7, 40.2] | 0.864 | 30.3<br>[24.4, 41.0] | 32.1<br>[25.5, 40.7] | 0.317 | 29.0<br>[23.9, 38.4] | 28.2<br>[23.2, 34.9] | 0.243 | **0.004** | **<0.001** |
| **Education** (years) | 15.0<br>[12.0, 16.0] | 15.0<br>[12.0, 17.0] | 0.663 | 15.0<br>[12.3, 16.5] | 14.5<br>[13.0, 16.0] | 0.823 | 13.0<br>[12.0, 16.0] | 13.0<br>[12.0, 15.0] | 0.323 | **<0.001** | **<0.001** |
| **BMI** (kg/m<sup>2</sup>) | 23.3<br>[21.4, 26.3] | 24.7<br>[23.2, 27.3] | **<0.001** | 24.0<br>[21.8, 27.2] | 26.0<br>[22.9, 28.1] | **0.004** | 23.9<br>[21.6, 27.2] | 25.8<br>[23.1, 29.3] | **<0.001** | 0.271 | 0.029 |
| **IQ** | 112.0<br>[107.0, 119.0] | 115.0<br>[108.8, 120.0] | **0.006** | 108.0<br>[101.0, 116.0] | 109.0<br>[103.0, 117.0] | 0.347 | 104.0<br>[95.0, 112.0] | 107.0<br>[94.3, 114.0] | 0.283 | **<0.001** | **<0.001** |
| **Age of Onset** (years) |  |  |  | 18.0<br>[15.0, 23.0] | 19.0<br>[16.0, 26.8] | **0.018** | 22.0<br>[18.0, 27.5] | 22.0<br>[19.0, 27.0] | 0.489 | **<0.001** | **0.003** |
| **DOI** (years) |  |  |  | 11.0<br>[6.4, 17.9] | 9.8<br>[5.4, 16.8] | 0.203 | 5.7<br>[2.3, 11.5] | 4.4<br>[1.6, 9.1] | **0.011** | **<0.001** | **<0.001** |
| **GAF, symptom** |  |  |  | 60.0<br>[51.0, 67.0] | 60.0<br>[52.0, 65.0] | 0.932 | 45.0<br>[38.0, 55.0] | 41.0<br>[37.0, 51.0] | **0.025** | **<0.001** | **<0.001** |
| **GAF, function** |  |  |  | 56.5<br>[48.0, 65.0] | 54.0<br>[47.0, 67.0] | 0.654 | 45.0<br>[40.0, 55.0] | 44.0<br>[38.0, 52.0] | **0.044** | **<0.001** | **<0.001** |
| **PANSS, total** |  |  |  | 43.0<br>[37.0, 49.0] | 43.0<br>[38.3, 50.0] | 0.333 | 54.5<br>[44.0, 66.3] | 60.0<br>[51.0, 71.0] | **<0.001** | **<0.001** | **<0.001** |
| **PANSS, negative** |  |  |  | 9.0<br>[7.0, 11.0] | 10.0<br>[8.0, 12.0] | **0.008** | 12.0<br>[9.0, 17.0] | 15.0<br>[11.0, 20.0] | **<0.001** | **<0.001** | **<0.001** |
| **PANSS, positive** |  |  |  | 9.0<br>[7.0, 10.0] | 9.0<br>[7.0, 11.0] | 0.569 | 13.0<br>[9.0, 16.0] | 14.0<br>[11.0, 17.0] | **0.019** | **<0.001** | **<0.001** |
| **YMRS** |  |  |  | 2.0<br>[0.0, 4.8] | 2.0<br>[0.0, 4.0] | 0.964 | 1.0<br>[0.0, 5.0] | 2.5<br>[0.0, 8.0] | **0.021** | 0.611 | 0.054 |
| **IDS** |  |  |  | 16.0<br>[9.0, 26.0] | 13.0<br>[7.0, 20.3] | **0.014** | 15.0<br>[7.0, 25.0] | 15.0<br>[7.0, 25.0] | 0.699 | 0.180 | 0.154 |
| **Antipsychotics**, user N (%) |  |  |  | 113 (61.7) | 96 (73.3) | **0.044** | 171 (92.4) | 259 (97.0) | **0.046** | **<0.001** | **<0.001** |
| DDD |  |  |  | 0.5<br>[0.3, 1.0] | 0.9<br>[0.5, 1.3] | **0.007** | 1.0<br>[0.7, 1.8] | 1.2<br>[0.8, 1.8] | 0.296 | **<0.001** | **<0.001** |
| **Antidepressants**, user N (%) |  |  |  | 77 (41.6) | 34 (26.0) | **0.006** | 65 (35.1) | 74 (27.7) | 0.115 | **<0.001** | **<0.001** |
| DDD |  |  |  | 1.0<br>[1.0, 2.0] | 1.5<br>[1.0, 2.0] | 0.265 | 1.0<br>[1.0, 1.6] | 1.5<br>[1.0, 2.0] | 0.113 | 0.695 | 0.828 |
| **Lithium**, user N (%) |  |  |  | 35 (18.9) | 20 (15.3) | 0.488 | 6 (3.2) | 5 (1.9) | 0.536 | **<0.001** | **<0.001** |
| DDD |  |  |  | 1.0<br>[1.0, 1.3] | 1.0<br>[0.8, 1.4] | 0.918 | 0.6<br>[0.5, 0.8] | 0.8<br>[0.5, 1.0] | 0.400 | **0.002** | 0.121 |
| **Antiepileptics**, user N (%) |  |  |  | 71 (38.4) | 44 (33.6) | 0.451 | 29 (15.7) | 29 (10.9) | 0.173 | **<0.001** | **<0.001** |
| DDD |  |  |  | 0.7<br>[0.3, 1.0] | 0.8<br>[0.6, 1.1] | 0.150 | 0.7<br>[0.6, 0.8] | 0.7<br>[0.5, 0.9] | 0.660 | 0.924 | 0.242 |
| **ICV** (liter) | 1.5<br>[1.4, 1.5] | 1.6<br>[1.5, 1.7] | **<0.001** | 1.5<br>[1.4, 1.6] | 1.7<br>[1.6, 1.8] | **<0.001** | 1.5<br>[1.4, 1.5] | 1.6<br>[1.5, 1.8] | **<0.001** | 0.072 | 0.117 |

\*Non-normal continuous data in median [Interquartile range] and categorical data as number %. Abbreviations: CTL = healthy controls, BD = bipolar disorders, SZ = schizophrenia disorders, N = number, R = right, L = left, A = ambidextrous, M = missing, y = year, BMI = body mass index, IQ = intelligence quotient, GAF = global assessment of functioning, PANSS = positive and negative syndrome scale, YMRS = young mania rating scale, IDS = inventory for depressive symptomatology, DDD = defined daily dose, ICV = intracranial volume, Dx = Diagnosis. Significant results are highlighted in bold (based on Chi<sup>2</sup> test for categorical data and Kruskal-Wallis test for continuous data).

To merge SZ and BD test scores from both batteries for each cognitive domain, we computed z-scores for both sexes combined based on the performance of the healthy control group. Z-scores were calculated using the following formula:

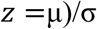

where X is the raw cognitive score, μ is the mean and σ is standard deviation of the healthy control group.

### Medication use

In the clinical groups, current use of the following medication was recorded and converted into Defined Daily Dose (DDD) ^38^: antipsychotics, antidepressants, and antiepileptics. In individuals with BD, lithium user status (yes/no) and serum concentration were also assessed (details published elsewhere ^39^).

### Neuroimaging data acquisition

T1-weighted images were acquired either at 1.5T (2004-2009, N = 745) or at 3T (2011-2019, N = 776). At 1.5T, images were acquired on a Siemens Magnetom Sonata scanner. At 3T, participants were scanned either on General Electric Signa HDxt scanner (n = 438) or a General Electric Discovery 750 scanner (n = 338). For details about the MRI acquisition parameters, see supplementary materials Note 2.

### Neuroimaging data processing

The FreeSurfer software package (version 6.0.0) was used to process T1-weighted images to extract volume estimates for left and right hippocampal total and subfield volumes as well as intracranial volume (ICV); estimated based on the Talairach transform. The automated segmentation of the hippocampus is based on a probabilistic atlas, created with ultra-high-resolution *ex vivo* MRI data (∼ 0.1–0.20 mm isotropic). It includes 12 subfields: cornu ammonis (CA) 1, CA3, CA4, fimbria, granule cell layer of dentate gyrus (GC-ML-DG), hippocampus-amygdala-transition-area (HATA), hippocampal tail, hippocampal fissure, molecular layer, parasubiculum, presubiculum and subiculum. Segmented images were quality controlled, and if necessary, manual editing of the surface reconstruction was performed by trained assistants following standard FreeSurfer procedures ^40^. We used ComBat, a batch adjustment tool adopted from the genomics literature, to remove unwanted variation associated with scanner whilst preserving biological associations in the data ^41, 42^. ComBat harmonization was performed on total hippocampal and subfield volumes as well as ICV, with empirical Bayes to leverage information across volumes, and with age, sex and diagnostic group as biological variables of interest. Hippocampal volumes before and after ComBat-harmonization are visualized in the supplementary materials (Figure S1-S2).

### Statistical analysis

All statistical tests were conducted in R, v4.2.2. False discovery rate (FDR) correction ^43^ was applied to the main analyses and sets of medication, clinical and cognitive analyses, separately, to account for multiple comparisons. All analyses included combined hippocampal volumes across both hemispheres (left + right volume) as dependent variables. Results from the main analyses for the left and right hippocampal subfields are reported in supplementary materials Table S1. Continuous variables in interaction terms were standardized (subtracting the mean and dividing by the SD) before the regression analysis.

### Sex and diagnostic group differences in hippocampal volume

To examine sex-by-diagnostic group differences in hippocampal volumes, we used analysis of covariance (ANCOVA, type III). Total hippocampal volume and each subfield volume were entered as dependent variables in separate models, which included diagnostic group-by-sex interaction as a fixed factor and age, age^2^, and ICV as covariates. Age^2^ was added to more accurately model the effect of age, which may have a nonlinear relationship with volume ^44^. Levene’s tests (volume∼diagnostic group) were performed to test whether the key assumption of ANCOVAs – homogeneity of variance – was met. No violations were detected. *Post hoc* Tukey tests were performed to contrast volume differences between females vs. males, females vs. females and males vs. males for each diagnostic group. Effect sizes were calculated as partial eta-squared (partial η^2^) based on the F-statistic. Based on diagnostic plots, one outlier was removed from the analysis (male with bipolar I disorder, Cook’s distance > 0.5). Case-control differences in hippocampal volumes, corrected by sex, age, age^2^, and ICV, are reported in Table S2 in the supplemental material. To explore the absolute differences in hippocampal volumes by sex and diagnostic group, we re-ran the main analyses without adjustment for ICV.

To further evaluate hippocampal volume differences between diagnostic groups across age, we fitted multiple regression models with total hippocampal and subfield volumes as dependent variables and group-by-age interaction, group-by-age^2^ interaction, sex and ICV as independent variables. Additional multiple regression models were fitted to further examine sex-by-diagnostic subgroup differences (i.e., bipolar I, bipolar II, SCZ-AF, SCZ, OPD) in total hippocampal and subfield volumes, adjusting for age, age^2^ and ICV. Due to sample size limitations, individuals with schizophreniform disorder or bipolar not otherwise specified were excluded from the diagnostic subgroup analysis. Effect sizes for multiple regression results were calculated as Cohen’s d based on the t-statistic.

### Sex-specific associations between medication and hippocampal volumes

To test if putative associations between medication use and hippocampal volumes (dependent variable) differed between the sexes, additional multiple regression models were fitted using an interaction term (sex-by-DDD) in SZ and BD separately. In both SZ and BD, the effects of antipsychotics, antidepressants, and antiepileptics were assessed as DDD (independent variable). In BD, we further explored the effects of lithium use status (yes/no) and serum concentration levels, on hippocampal volumes, in separate multiple regression models. All models were adjusted for age, age^2^, and ICV.

To investigate the sex-adjusted main effects of the aforementioned independent variables (antipsychotics, antidepressants, antiepileptics, lithium) on hippocampal volumes, we fitted separate regression models for BD and SZ, adjusting for sex, age, age^2^, and ICV.

### Sex-specific associations between clinical measures and hippocampal volumes

The aforementioned model specifications were used to examine the associations between hippocampal volumes and psychotic symptoms (PANSS, negative/positive-subscale) in SZ, affective symptoms (YMRS and IDS) in BD, age of onset, duration of illness, and general functioning (GAF symptoms/function) between the sexes. As both age of onset (BD r = 0.58, SZ r = 0.66) and duration of illness (BD r = 0.66, SZ r = 0.62) had high Pearson correlations with age, these linear models were only adjusted for ICV, whereas the other models were adjusted for age, age^2^ and ICV.

To assess the sex-adjusted main effects of the before mentioned independent variables on hippocampal volumes, we fitted separate regression models for BD and SZ, adjusting for sex, age, age^2^ and ICV. The age of onset and duration of illness models were again only adjusted for sex and ICV.

### Sex-specific associations between cognitive measures and hippocampal volumes

To investigate associations between cognitive measures (verbal learning, verbal memory, processing speed, working memory) and hippocampal volumes between females and males, regression models with a sex-by-cognitive measure interaction term were fitted. These models were adjusted for age, age^2^, ICV and years of education.

The sex-adjusted main effects of cognitive measures on hippocampal volumes were tested in separate models, covarying for the beforementioned covariates and sex.

## Results

### Demographic and clinical variables

Sample demographics and clinical characteristics, stratified by sex and diagnostic group, are summarized in Table 1. Cognitive measures, stratified by diagnostic group and sex, are displayed in Table 2.

**Table 2.** Overview of neurocognitive domains and corresponding tests, stratified by diagnostic group and sex.

|  | BD |  |  | SZ |  |  |
| --- | --- | --- | --- | --- | --- | --- |
|  | Female | Male | p | Female | Male | p |
| N | 173 | 126 |  | 172 | 250 |  |
| **Cognitive measures, z-scores** |  |  |  |  |  |  |
| **Verbal learning** | 0.16 ± 0.95 | -0.45 ± 1.31 | **<0.001** | -0.62 ± 1.25 | -0.97 ± 1.22 | **0.005** |
| CVLT, list A total correct | 0.20 ± 0.94 | -0.48 ± 1.38 | **<0.001** | -0.49 ± 1.13 | -0.92 ± 1.20 | **0.004** |
| HVLT-R, immediate recall | 0.08 ± 0.97 | -0.38 ± 1.18 | **0.037** | -0.83 ± 1.41 | -1.05 ± 1.27 | 0.306 |
| **Verbal memory** | 0.11 ± 0.94 | -0.57 ± 1.37 | **<0.001** | -0.77 ± 1.34 | -0.93 ± 1.32 | 0.244 |
| CVLT, long delay free recall | 0.12 ± 0.96 | -0.65 ± 1.45 | **<0.001** | -0.55 ± 1.20 | -0.87 ± 1.27 | **0.042** |
| HVLT-R, delayed recall | 0.07 ± 0.90 | -0.42 ± 1.16 | **0.022** | -1.21 ± 1.51 | -1.05 ± 1.41 | 0.540 |
| **Processing speed** | -0.44 ± 1.15 | -0.90 ± 1.11 | **0.001** | -0.97 ± 1.16 | -1.39 ± 1.21 | **<0.001** |
| Digit Symbol Coding Test from WAIS-III | -0.55 ± 1.20 | -0.89 ± 1.12 | **0.044** | -0.99 ± 1.10 | -1.29 ± 1.19 | **0.038** |
| Brief Assessment of Cognition in Schizophrenia | -0.23 ± 1.02 | -0.93 ± 1.12 | **0.002** | -0.93 ± 1.27 | -1.56 ± 1.23 | **0.002** |
| **Working memory** | -0.51 ± 0.95 | -0.54 ± 0.94 | 0.787 | -0.82 ± 0.99 | -0.77 ± 1.01 | 0.625 |
| Letter-Number Sequencing Test from the D-KEFS battery | -0.64 ± 0.98 | -0.65 ± 0.93 | 0.951 | -0.89 ± 0.99 | -0.80 ± 1.03 | 0.517 |
| Letter-Number Sequencing Test from the MCCB | -0.28 ± 0.86 | -0.35 ± 0.95 | 0.687 | -0.69 ± 0.99 | -0.71 ± 0.97 | 0.946 |

Mean ± standard deviation. Abbreviation: BD = bipolar disorders, SZ = schizophrenia spectrum disorders, N = sample size, CVLT = California verbal learning test, HVLT-R = Hopkins verbal learning test revised, WAIS = Wechsler adult intelligence scale, D-KEFS = Delis-Kaplan executive function system, MCCB = MATRICS consensus cognitive battery. Significant results are highlighted in bold (Kruskal-Wallis test for continuous data).

**Table 3.** Sex, diagnostic group and sex-by-diagnostic group effects on total hippocampal and subfield volumes assessed using analysis of covariance.

| Subfield | Variable | F-value | partial $\eta^2$ | p-value | p<sub>FDR</sub>-value |
| --- | --- | --- | --- | --- | --- |
| CA1 | Diagnostic group | 8.575 | 0.011 | **1.98e-04** | **0.001** |
|  | Sex | 16.114 | 0.011 | **6.26e-05** | **0.000** |
|  | Group-by-sex interaction | 2.334 | 0.003 | 0.097 | 0.119 |
| CA3 | Diagnostic group | 3.098 | 0.004 | **0.045** | 0.066 |
|  | Sex | 3.704 | 0.002 | 0.054 | 0.071 |
|  | Group-by-sex interaction | 3.126 | 0.004 | **0.044** | 0.066 |
| CA4 | Diagnostic group | 5.237 | 0.007 | **0.005** | **0.012** |
|  | Sex | 3.853 | 0.003 | 0.050 | 0.069 |
|  | Group-by-sex interaction | 4.944 | 0.006 | **0.007** | **0.016** |
| Fimbria | Diagnostic group | 0.103 | 1.36e-04 | 0.902 | 0.902 |
|  | Sex | 23.009 | 0.015 | **1.77e-06** | **3.45e-05** |
|  | Group-by-sex interaction | 3.914 | 0.005 | **0.020** | **0.033** |
| GC-ML-DG | Diagnostic group | 6.666 | 0.009 | **0.001** | **0.005** |
|  | Sex | 6.455 | 0.004 | **0.011** | **0.021** |
|  | Group-by-sex interaction | 5.866 | 0.008 | **0.003** | **0.008** |
| HATA | Diagnostic group | 8.937 | 0.012 | **1.39e-04** | **0.001** |
|  | Sex | 15.237 | 0.010 | **9.90e-05** | **0.001** |
|  | Group-by-sex interaction | 4.012 | 0.005 | **0.018** | **0.031** |
| Hippocampal tail | Diagnostic group | 1.584 | 0.002 | 0.205 | 0.229 |
|  | Sex | 9.354 | 0.006 | **0.002** | **0.006** |
|  | Group-by-sex interaction | 2.298 | 0.003 | 0.101 | 0.119 |
| Hippocampal fissure | Diagnostic group | 1.899 | 0.003 | 0.150 | 0.172 |
|  | Sex | 28.125 | 0.018 | **1.31e-07** | **5.09e-06** |
|  | Group-by-sex interaction | 0.188 | 2.49e-04 | 0.828 | 0.850 |
| Molecular layer | Diagnostic group | 8.118 | 0.011 | **3.11e-04** | **0.001** |
|  | Sex | 8.679 | 0.006 | **0.003** | **0.008** |
|  | Group-by-sex interaction | 4.749 | 0.006 | **0.009** | **0.018** |
| Parasubiculum | Diagnostic group | 0.696 | 0.001 | 0.499 | 0.526 |
|  | Sex | 11.692 | 0.008 | **0.001** | **0.003** |
|  | Group-by-sex interaction | 2.921 | 0.004 | 0.054 | 0.071 |
| Presubiculum | Diagnostic group | 2.561 | 0.003 | 0.078 | 0.098 |
|  | Sex | 17.224 | 0.011 | **3.51e-05** | **3.42e-04** |
|  | Group-by-sex interaction | 4.092 | 0.005 | **0.017** | **0.030** |
| Subiculum | Diagnostic group | 1.169 | 0.002 | 0.311 | 0.337 |
|  | Sex | 6.708 | 0.004 | **0.010** | **0.019** |
|  | Group-by-sex interaction | 3.266 | 0.004 | **0.038** | 0.060 |
| Hippocampus | Diagnostic group | 6.608 | 0.009 | **0.001** | **0.005** |
|  | Sex | 19.027 | 0.012 | **1.38e-05** | **1.79e-04** |
|  | Group-by-sex interaction | 6.114 | 0.008 | **0.002** | **0.006** |

The statistical results are based on analysis of covariance (adjusted for age, age<sup>2</sup>, intracranial volume). Abbreviation: CA = cornu ammonis, GC-ML-DG = granule cells in the molecular layer of the dentate gyrus, HATA = hippocampal-amygdaloid transition area, FDR = false discovery rate. Significant results are highlighted in bold.

### Sex and diagnostic group differences in hippocampal volumes

In the main models, including a sex-by-diagnostic group interaction term, total hippocampal, CA1, CA4, GC-ML-DG, HATA and molecular layer volumes were significantly smaller in individuals with SZ and BD relative to healthy controls (see Table 2, Figure 1). All hippocampal volumes were smaller in SZ and BD relative to healthy controls, when not accounting for potential sex-by-diagnostic group interactions (see Table S2).

**Figure 1.**
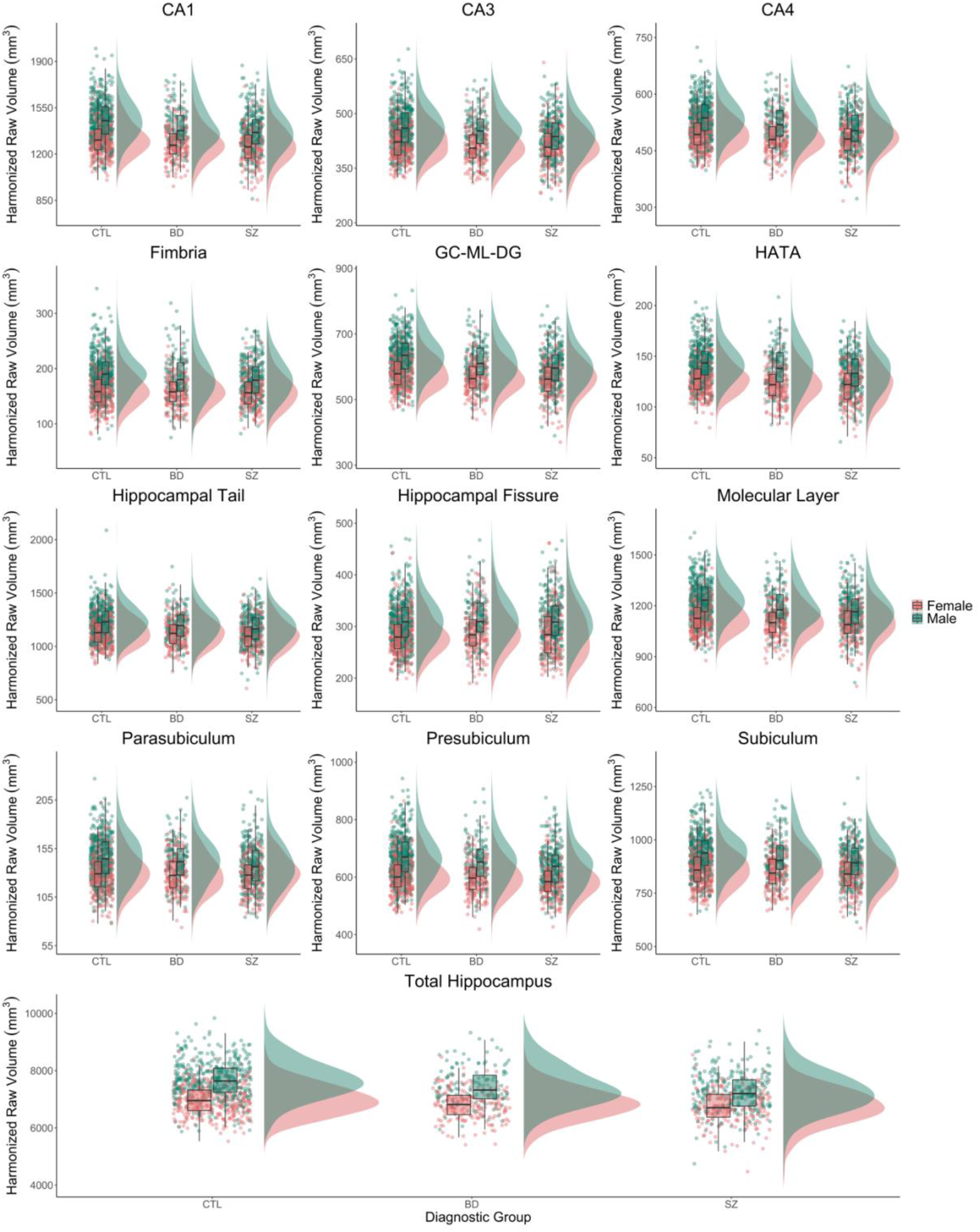
Hippocampal volumes stratified by diagnostic group and sex. ComBat-harmonized volumetric data is displayed as raincloud plots, which combines boxplots, unadjusted raw data points (scatterplot), and the distributions of the data (histogram) using split-half violins. CTL = healthy controls, BD = bipolar disorders, SZ = schizophrenia spectrum disorders, CA = cornu ammonis, GC-ML-DG = granule cells in the molecular layer of the dentate gyrus, HATA = hippocampal-amygdaloid transition area.

We found significant main effects of sex for the total hippocampus and for 10 of the 12 hippocampal subfields, not including CA3 and CA4. Significant sex-by-group interactions were found for the total hippocampus, CA4, fimbria, GC-ML-DG, HATA, presubiculum, and molecular layer. Effect sizes were overall small (partial η^2^ ≤ 0.018). When not adjusting for ICV, we found significant sex-by-group interactions for total hippocampus volume and GC-ML-DG after FDR-correction (see Table S3). Post hoc Tukey tests revealed higher volumes in males than females in the healthy control group in most subfields, but not in BD or SZ (details for subfields, see supplementary material Table S4). We further found significantly higher total hippocampus volumes in males with BD relative to males with SCZ, which was likely driven by significantly higher volumes in the HATA and GC-ML-DG, and nominally higher volumes in CA4 (Table S4). No volumetric differences were found between females with BD and SCZ.

Age and age^2^ showed a significant main effect on all volumes, except the presubiculum and parasubiculum, based on the ANCOVA models with sex-by-group interactions. Before FDR-correction, we found a significant group-by-age and group-by-age^2^ interaction in individuals with SZ relative to healthy controls for the presubiculum (age: t = -1.943, p = 0.052, p_FDR_ = 0.657; age^2^: t = 2.090, p = 0.037, p_FDR_ = 0.638) and parasubiculum (age: t = -3.010, p = 0.003, p_FDR_ = 0.069; age^2^: t = 3.223, p = 0.001, p_FDR_ = 0.067) in the regression models.

The diagnostic subgroup analysis revealed significant sex-by-group effects for GC-ML-DG, molecular layer, fimbria, CA4 and total hippocampus volumes in SCZ only (Figure 2, supplementary materials Table S5).

**Figure 2.**
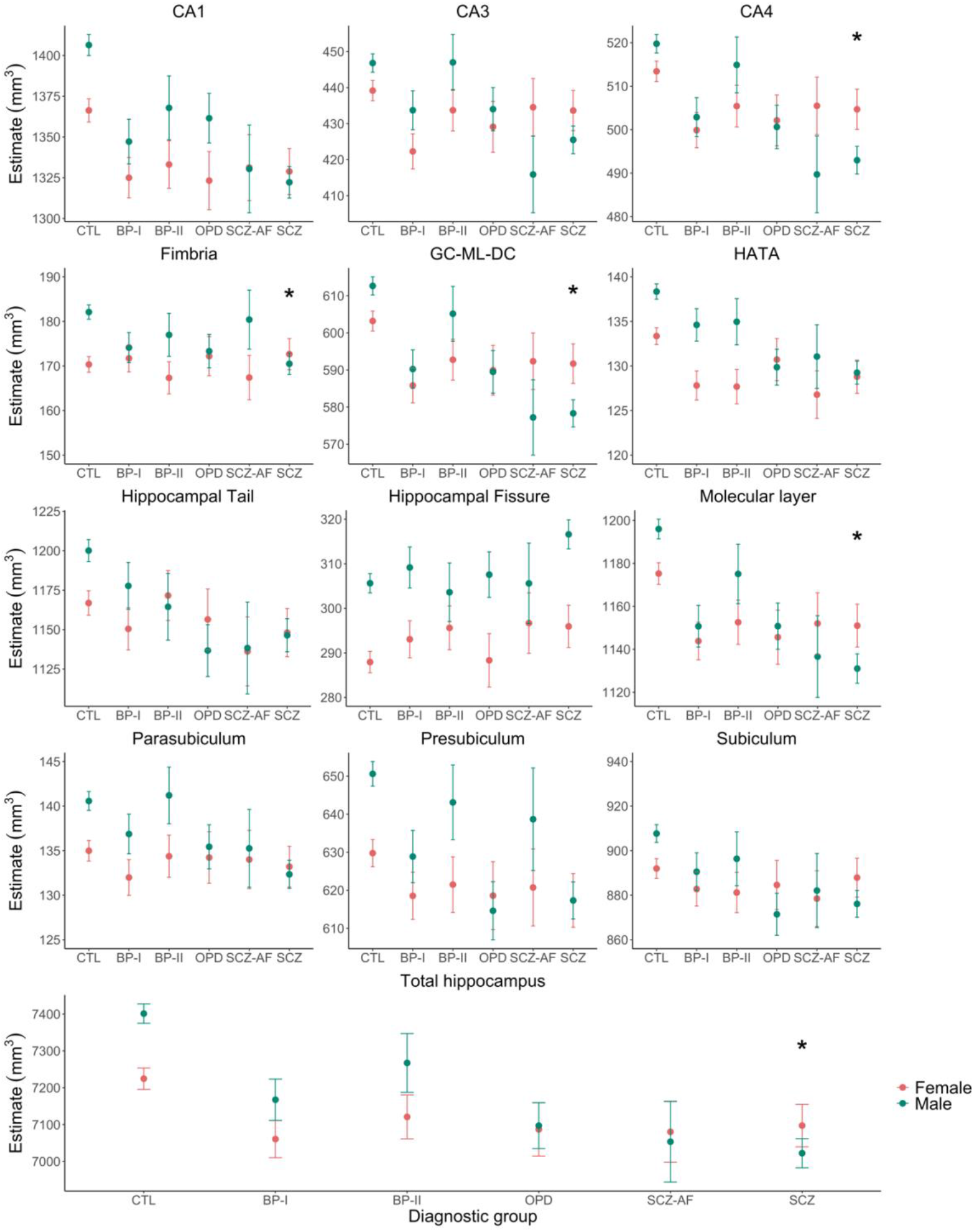
Estimates of total hippocampus and subfield volumes stratified by diagnostic subgroup and sex. The model is adjusted for age, age^2^, and intracranial volume. Estimates are displayed with upper and lower confidence intervals. Stars represent significant group difference relative to healthy controls (CTL), after false discovery rate correction. Significance codes: p < 0.01 ‘*’. Abbreviation: B = bipolar, SCZ = schizophrenia, SCZ-AF = schizoaffective disorder, OPD = other psychotic disorders, CA = cornu ammonis, GC-ML-DG = granule cells in the molecular layer of the dentate gyrus, HATA = hippocampal-amygdaloid transition area.

### Sex-specific associations between medication use and hippocampal volumes

After FDR-correction, we found no significant main effects or sex-by-medication interaction effects between antipsychotic, antidepressant, antiepileptic medication use (DDD) and total hippocampus and subfield volumes in SZ and BD (see Table S7).

### Sex-specific associations between clinical characteristics and hippocampal volumes

We found a significant main effect for the association between fissure volumes and age of onset (t = 4.149, p = 4.32e-05, p_FDR_ = 0.007) as well as duration of illness (t = 3.713, p = 2.43e-04, p_FDR_ = 0.025) in BD. Before FDR-correction, we also found a significant interaction by sex for both variables of interest (age of onset: t = 2.272, p = 0.024, p_FDR_ = 0.531; duration of illness: t = -2.325, p = 0.021, p_FDR_ = 0.498). Similarly, in SZ, we also found a significant association between age of onset and duration of illness and fissure volume. Only the association with duration of illness was significant after FDR-correction (t = 4.432, p = 1.18e-05, p_FDR_ = 0.004). No other main or interaction effects survived FDR-correction. All associations are detailed in Table S8.

### Sex-specific associations between cognitive measures and hippocampal volumes

We found no significant associations between cognitive measures (z-scores) and hippocampal volumes in SZ and BD, after FDR-correction (see Table S9).

## Discussion

Using a well-powered sample, our findings indicate that the pattern of morphological sex differences in hippocampal subfields is altered in individuals with schizophrenia relative to healthy controls, due to higher volumetric deficits in males. Effect sizes were overall small.

Alterations in hippocampal morphology in psychiatric illnesses have been widely reported, both on a macro- and microscopic level. Here, we found significantly lower total hippocampal, CA1, CA4, GC-ML-DG, molecular layer and HATA volumes in individuals with schizophrenia spectrum disorder (SZ) and bipolar disorders (BD) relative to healthy controls, when accounting for sex-by-diagnostic group interactions and ICV. When not adjusting for this interaction, all 12 hippocampal subfields were smaller in SZ and BP relative to healthy controls, which is in line with previous results ^20^. When not accounting for ICV, significant sex-by-group interactions for total hippocampus volume and the GC-ML-DG remained after FDR-correction.

While diagnostic group differences in hippocampal subfield volumes have been extensively demonstrated, sex differences in hippocampal volumes are still debated. In healthy controls, region-specific sex differences in hippocampal subfield volumes have been demonstrated, with larger volumes in males relative to females, most prominently in the fimbria and parasubiculum ^23^. In healthy controls, our post hoc Tukey pair-wise comparison revealed significantly higher volumes in males relative to females for the total hippocampus, parasubiculum, presubiculum, CA1, molecular layer, HATA, fimbria, hippocampal tail and fissure, after adjustment for intracranial volume (ICV). We found no evidence for larger volumes in healthy female controls as compared to males for any of the subfields. However, it should be noted that sex differences in neuroanatomical structure may be dependent on ICV estimation ^45^ and choice of statistical method for adjusting for ICV ^46-48^. Here, we estimated ICV as estimated total ICV via Freesurfer v6.0.0 and used the ANCOVA method, which has been shown to more effectively remove ICV-related variation than the proportions method ^46^. While we found higher volumes for most subfields in healthy males relative to females, this sex difference was largely absent or even reversed in individuals with SZ and BD. Directly comparing SZ and BD, we found lower total hippocampal, GC-ML-DG and HATA volumes in males with SZ relative to males with BD. No differences were found between females with BD and SZ. This finding is in line with previous studies showing greater magnitudes of hippocampal subfield volume reductions in schizophrenia than in bipolar disorder ^12, 22^. However, our results suggest that this differential pattern of volume deficits is not just dependent on diagnosis but also on sex.

In a previous study using the same study sample, we found no significant group-by-sex interaction for bilateral total amygdala volume and amygdala nuclei volumes ^39^, suggesting that sex-by-diagnostic group differences may be specific to the hippocampus within the amygdala-hippocampus formation. The diagnostic subgroup analysis revealed significant sex-by-group effect on hippocampal volume in SCZ only, particularly for the GC-ML-DG, molecular layer, fimbria, CA4 and total hippocampus. In these subfields, volumes were significantly lower in males with schizophrenia as compared with healthy controls, but not in females with schizophrenia. This finding is in line with previous studies, showing lower total hippocampal or medial temporal lobe volume in males with schizophrenia, but not in females with schizophrenia relative to same sex healthy controls ^49, 50^.

Our findings suggest that this male-specific deficit in total hippocampal volume may be driven by lower volume in the CA4, GC-ML-DG, molecular layer and fimbria. Previous postmortem studies found significantly lower oligodendrocyte number in the CA4 ^51^, and decreased stem cell proliferation ^52^ as well as neuron numbers in the DG ^53^, suggesting disturbed hippocampal connectivity and impaired adult neurogenesis in schizophrenia. Sex-specific effects in these studies are either inconclusive, due to small sample sizes, or not studied. The CA4 is considered a starting point for axonal fibers of the fornix, and fornix fibers traverse through the fimbria, which connect the hippocampus with several cortical and subcortical regions. As the hippocampus and fornix are anatomically tightly connected, lower CA4 and fimbria volume may contribute to impaired fornix integrity, reported in schizophrenia ^54^. Interestingly, impaired fornix-hippocampus integrity has previously been linked both to early psychosis ^55^ and cognitive disturbances and memory function in schizophrenia ^56^. Furthermore, the molecular layer has also been linked to memory function ^57^. One might speculate that the observed male-specific volume reductions in the CA4, GC-ML-DG, fimbria and molecular layer in schizophrenia may be linked to cognitive impairments, which have been shown to be more pronounced in males than females with schizophrenia ^28^. Even though we found sex differences in SZ on verbal learning, verbal memory and processing speed tasks, where females out performed males, we did not find any significant interaction effects between cognitive measures and volumes in the aforementioned subfields by sex, after FDR-correction. Similarly, none of the observed main effects of cognitive measures on subfield-specific volumes survived correction for multiple comparisons. This finding is not in line with previous results ^13, 58, 59^. However, previous studies have either focused on a few cognitive tests ^13, 58^, not multiple domains, or have studied the whole hippocampus and not hippocampal subfields ^59^, reducing the number of multiple comparisons. Furthermore, while we selected tasks that have previously been linked to hippocampal volumes and have been shown to be impaired in SZ and BD, these cognitive tasks are not specific to the hippocampus; but also rely on other brain regions such as the dorsolateral prefrontal cortex ^60^. More research, including cognitive batteries particularly sensitive to hippocampal functioning, is needed to elucidate the relationship between cognitive functioning, hippocampal morphology and sex in SZ and BD.

Although medication, including antipsychotics ^61^, antidepressants ^62^ and lithium ^63^ have been associated with hippocampal volumes, we found no association between current use of psychotropic drugs or mood stabilizers and volumes in SZ and BD after FDR-correction. However, as medication effects might depend on the specific types of drugs and duration of exposure which may not necessarily be linear, the use of DDD alone to address this confound has limitations. Similarly, we found no associations between most clinical measures (i.e., GAF scores, PANSS, negative/positive-subscale in SZ, and YMRS and IDS in BD) and hippocampal volumes after FDR-correction. The lack of an association between psychotic symptoms and volumes in SZ may result from the relatively small variation in psychotic symptoms in our sample. It may also relate to issues such as variability of symptom states over time, or currently unknown confounders. We did, however, find a significant main effect of age of onset and duration of illness on hippocampal fissure volume in BD and SZ, suggesting a positive association. The hippocampal fissure does not represent hippocampal tissue, but rather a cerebral spinal fluid cleft that forms during early brain development ^64^. The presence of enlarged hippocampal fissures has been shown in first-episode individuals with schizophrenia, and may be a sign of disrupted hippocampal development in schizophrenia ^65^.

A major strength of the current study is the large sample size with a balanced sex distribution, and detailed clinical assessment of individuals with schizophrenia spectrum disorders and bipolar disorders. However, the cross-sectional nature of the data precludes causal inferences, and longitudinal studies are needed to determine the timing of changes in hippocampal subfields by diagnostic group and sex. Furthermore, automated hippocampal subfield segmentation using MRI is challenging. The segmentation method used here is based on ultra-high-resolution *ex vivo* MRI data which allows for the precise delineation of hippocampal subfields to determine tissue priors ^18^. In the current study, however, subfields are probabilistically labeled based on conventional MRI data with 1 mm isotropic resolution. As such, the volumes of subfields contained within the interior of the hippocampal formation must be interpreted with caution, and a replication of our results using high resolution MRI data is warranted. Furthermore, the reliability of automated volumetry is inversely associated with volume size ^66^. The volumes of smaller hippocampal subfields, such as the hippocampal fissure, fimbria, parasubiculum and HATA, may thus be less reliable. In addition, although automated volumetry has been shown to be generally reliable in multicenter MRI studies ^66^, and we successfully harmonized volumes and ICV across scanners using ComBat (see Figure S1 and S2), residual effects of scanner may still be present. Beside the effect of sex, we also found significant main effects of age and age^2^ on all volumes, except the presubiculum and parasubiculum. This finding is partly in line with a study in healthy controls, reporting age-related volume changes in all subfields, except in the subiculum complex (subiculum proper, presubiculum, parasubiculum) ^67 60^. However, before FDR-correction, we found a significant age-by-diagnostic group interaction for individuals with SZ relative to healthy controls in the presubiculum and parasubiculum, potentially suggesting abnormal aging processes in these subfields in SZ.

In summary, the pattern of morphological sex differences in the hippocampus appears to be altered in individuals with schizophrenia relative to healthy controls, due to higher volumetric deficits in males with schizophrenia. Our findings highlight that sex should be considered when studying individuals with psychiatric illnesses, not just as a covariate, which might mask potential sex differences, but as a discovery variable.

## Data Availability

All data produced in the present study are available upon reasonable request to the authors.

## Acknowledgments

This work was supported by the Research Council of Norway (223273, 250358, 286838), the South-Eastern Norway Regional Health Authority (2017097, 2019104, 2020020) and the K. G. Jebsen Foundation (SKGJ-MED-008).

