## Supplementary Materials for "Altered sex differences in hippocampal subfield volumes in schizophrenia"

**Abstract**

**Objective:** The hippocampus is a heterogenous brain structure that differs between the sexes and has been implicated in the pathophysiology of psychiatric illnesses. Here, we explored sex and diagnostic group differences in hippocampal subfield volumes, in individuals with schizophrenia spectrum disorder (SZ), bipolar disorders (BD) and healthy controls.

**Methods:** 1,521 participants underwent T1-weighted magnetic resonance imaging (SZ, n = 452, mean age 30.7 ± 9.2 [SD] years, males 59.1%; BD, n = 316, 33.7 ± 11.4, 41.5%; healthy controls, n = 753, 34.1 ± 9.1, 55.6%). Total hippocampal, subfield, and intracranial volumes were estimated with Freesurfer (v6.0.0). Analysis of covariance and multiple regression models were fitted to examine sex-by-diagnostic (sub)group interactions in volume. In SZ and BD, separately, associations between volumes and clinical as well as cognitive measures were examined between the sexes using regression models.

**Results:** Significant sex-by-group interactions were found for the total hippocampus, dentate gyrus, molecular layer, presubiculum, fimbria, HATA, and CA4, indicating a larger volumetric deficit in male patients relative to female patients when compared with same-sex healthy controls. Subgroup analyses revealed that this interaction was driven by males with schizophrenia. Effect sizes were overall small (partial η^2^ < 0.02). We found no significant sex differences in the associations between hippocampal volumes and clinical or cognitive measures in SZ and BD.

**Conclusions:** Using a well-powered sample, our findings indicate that the pattern of morphological sex differences in hippocampal subfields is altered in individuals with schizophrenia relative to healthy controls, due to higher volumetric deficits in males.

**Key words**: hippocampus, neuroimaging, schizophrenia, bipolar disorders, sex differences

***Note 1: Participant inclusion***

Participants, aged between 18-65 years, were excluded if they had an intelligence quotient below 70, a history of moderate or severe head trauma resulting in hospitalization, and neurological disorders or somatic illness which could affect brain structure and function. While patients diagnosed with substance abuse were included, control participants with substance misuse disorder or cannabis use within the last 3 months were excluded.

***Note 2: Neuroimaging data acquisition***

At 1.5 T, two sagittal T1-weighted magnetization prepared rapid acquisition gradient-echo (MPRAGE) volumes were acquired with the Siemens tfl3d1_ns pulse sequence, and averaged to improve signal-to-noise ratio. The acquisition parameters were as follows: echo time (TE) = 3.93 msec, repetition time (TR) = 2730 msec, inversion time (TI) = 1000 msec, flip angle = 7º, field of view = 24 cm, voxel size = 1.33 x 0.94 x 1 mm3, number of partitions = 160.

At 3T, the scanning parameters were as follow: (1) T1-weighted 3D Fast Spoiled Gradient Echo (FSPGR, from 2011-2015): TR/TE = 7.8 ms/min, TI = 450 msec, flip angle = 12°, FOV = 256×256 mm2, slice thickness = 1.2 mm, acquisition matrix = 256×192, reconstruction matrix = 256×256; (2) inversion recovery-prepared 3D gradient recalled echo (BRAVO, from 2015-present): TR/TE/TI 8.16 ms/3.18 ms/450ms, slice thickness = 1.2 mm, reconstruction matrix = 256×256, flip angle = 12°.

|  | **Healthy Controls** | | |  | | **BD** | |  | **SZ** | |  |
| --- | --- | --- | --- | --- | --- | --- | --- | --- | --- | --- | --- |
| Scanner (Sequence), N (%) | **Female**  **N = 334** | **Male**  **N = 419** | **p-value** | | **Female**  **N = 185** | | **Male**  **N = 131** | **p-value** | **Female**  **N = 185** | **Male**  **N = 267** | **p-value** |
| 1.5-T, MPRAGE | 128 (38.3) | 142 (33.9) | 0.302 | | 108 (58.4) | | 76 (58.0) | 0.904 | 118 (63.8) | 173 (64.8) | 0.846 |
| 3-T, FSPGR | 115 (34.4) | 166 (39.6) |  | | 32 (17.3) | | 25 (19.1) |  | 40 (21.6) | 60 (22.5) |  |
| 3-T, BRAVO | 91 (27.2) | 111 (26.5) |  | | 45 (24.3) | | 30 (22.9) |  | 27 (14.6) | 34 (12.7) |  |

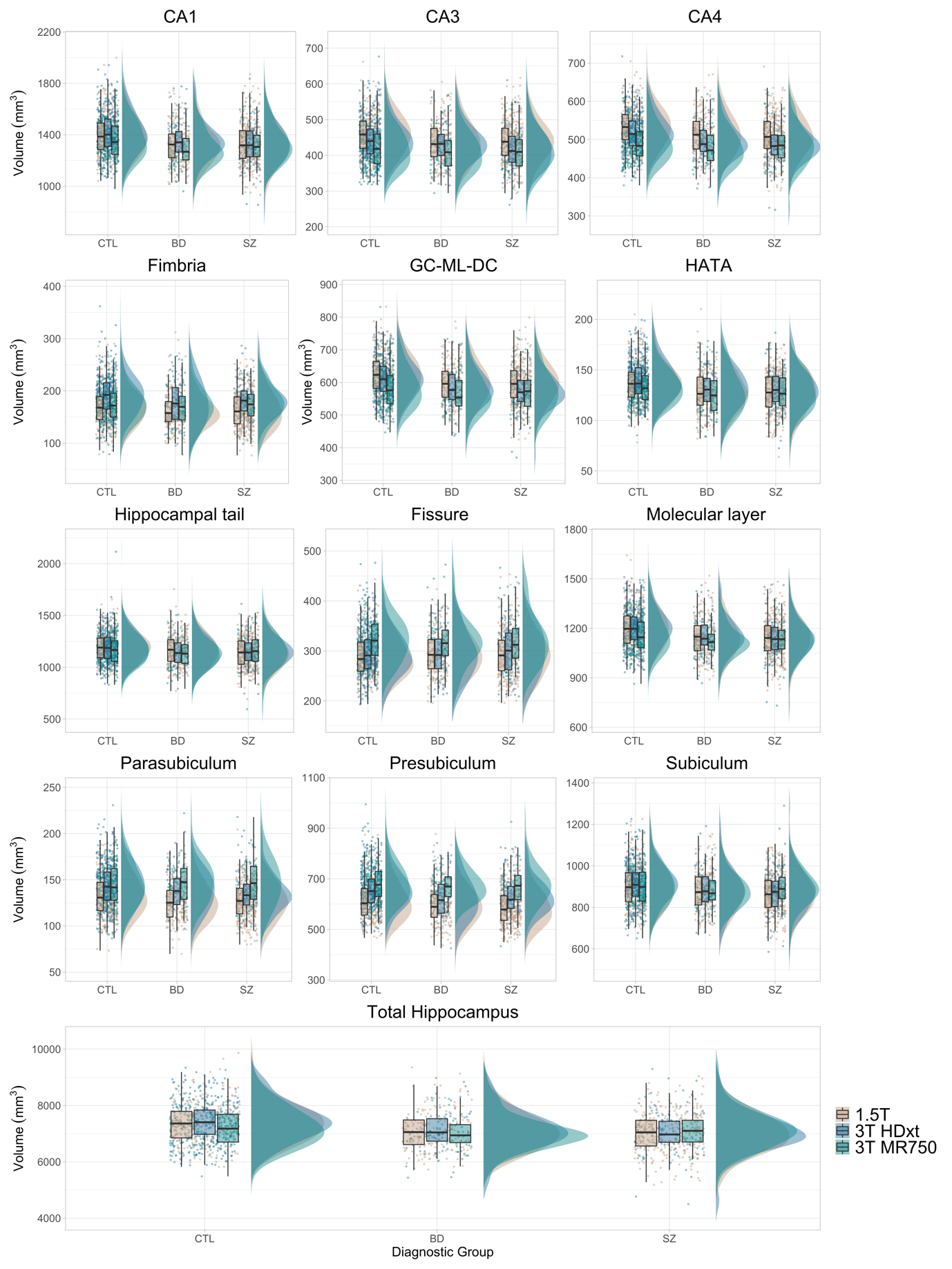

**Figure S1| Raw hippocampal volumes stratified by scanner.** The data is displayed as a raincloud plot, which combines boxplots, raw data points and the distribution of data using split-half violins. Abbreviations: CTL = healthy controls, BD = bipolar spectrum disorders, SZ = schizophrenia spectrum disorder.

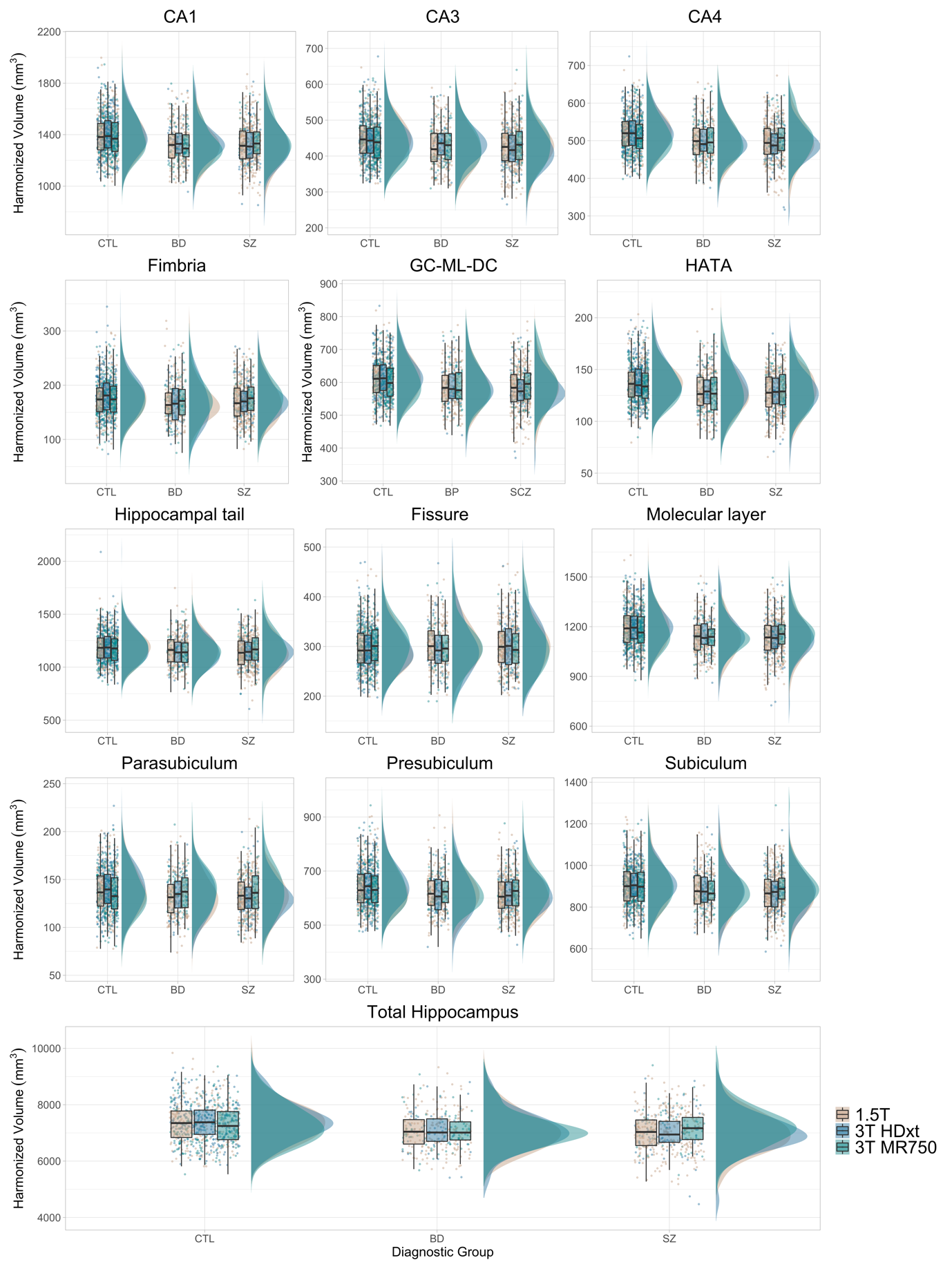

**Figure 2| ComBat-harmonized hippocampal volumes stratified by scanner.** The data is displayed as a raincloud plot, which combines boxplots, raw data points and the distribution of data using split-half violins. Abbreviations: CTL = healthy controls, BD = bipolar spectrum disorders, SZ = schizophrenia spectrum disorder.

**Table S1| Sex, diagnostic group and sex-by-diagnostic group effects on lateralized total hippocampal and subfield volumes assessed using analysis of covariance.**

| **Subfields** | **Side** | **Variable** | **F-value** | **partial η ^2^** | **p-value** | **p_FDR_-value** |
| --- | --- | --- | --- | --- | --- | --- |
| CA1 | L | Diagnostic group | 6.202 | 0.008 | **0.002** | **0.009** |
|  | L | Sex | 21.915 | 0.014 | **3.11e-06** | **6.08e-05** |
|  | L | Group-by-sex interaction | 2.915 | 0.004 | 0.055 | 0.082 |
|  | R | Diagnostic group | 8.357 | 0.011 | **2.46e-04** | **0.002** |
|  | R | Sex | 7.606 | 0.005 | **0.006** | **0.017** |
|  | R | Group-by-sex interaction | 1.265 | 0.002 | 0.282 | 0.319 |
| CA3 | L | Diagnostic group | 2.612 | 0.003 | 0.074 | 0.105 |
|  | L | Sex | 3.427 | 0.002 | 0.064 | 0.094 |
|  | L | Group-by-sex interaction | 3.292 | 0.004 | **0.037** | 0.062 |
|  | R | Diagnostic group | 2.582 | 0.003 | 0.076 | 0.105 |
|  | R | Sex | 2.517 | 0.002 | 0.113 | 0.149 |
|  | R | Group-by-sex interaction | 1.937 | 0.003 | 0.145 | 0.182 |
| CA4 | L | Diagnostic group | 4.171 | 0.005 | **0.016** | **0.032** |
|  | L | Sex | 4.121 | 0.003 | **0.043** | 0.068 |
|  | L | Group-by-sex interaction | 4.614 | 0.006 | **0.010** | **0.024** |
|  | R | Diagnostic group | 4.578 | 0.006 | **0.010** | **0.024** |
|  | R | Sex | 2.357 | 0.002 | 0.125 | 0.160 |
|  | R | Group-by-sex interaction | 3.625 | 0.005 | 0.027 | 0.049 |
| Fimbria | L | Diagnostic group | 0.155 | 0.000 | 0.856 | 0.868 |
|  | L | Sex | 12.662 | 0.008 | **3.85e-04** | **0.002** |
|  | L | Group-by-sex interaction | 2.742 | 0.004 | 0.065 | 0.094 |
|  | R | Diagnostic group | 0.181 | 0.000 | 0.834 | 0.868 |
|  | R | Sex | 24.930 | 0.016 | **6.63e-07** | **2.59e-05** |
|  | R | Group-by-sex interaction | 3.870 | 0.005 | **0.021** | **0.040** |
| GC-ML-DG | L | Diagnostic group | 5.299 | 0.007 | **0.005** | **0.016** |
|  | L | Sex | 5.904 | 0.004 | **0.015** | **0.032** |
|  | L | Group-by-sex interaction | 5.188 | 0.007 | **0.006** | **0.017** |
|  | R | Diagnostic group | 5.909 | 0.008 | **0.003** | **0.011** |
|  | R | Sex | 4.852 | 0.003 | **0.028** | **0.049** |
|  | R | Group-by-sex interaction | 4.591 | 0.006 | **0.010** | **0.024** |
| HATA | L | Diagnostic group | 8.324 | 0.011 | **2.54e-04** | **0.002** |
|  | L | Sex | 21.906 | 0.014 | **3.12e-06** | **6.08e-05** |
|  | L | Group-by-sex interaction | 3.506 | 0.005 | **0.030** | 0.052 |
|  | R | Diagnostic group | 6.238 | 0.008 | **0.002** | **0.009** |
|  | R | Sex | 5.737 | 0.004 | **0.017** | **0.033** |
|  | R | Group-by-sex interaction | 3.038 | 0.004 | **0.048** | 0.075 |
| Hippocampal fissure | L | Diagnostic group | 1.454 | 0.002 | 0.234 | 0.273 |
|  | L | Sex | 32.582 | 0.021 | **1.37e-08** | **1.07e-06** |
|  | L | Group-by-sex interaction | 0.298 | 0.000 | 0.742 | 0.782 |
|  | R | Diagnostic group | 1.694 | 0.002 | 0.184 | 0.224 |
|  | R | Sex | 13.773 | 0.009 | **2.14e-04** | **0.002** |
|  | R | Group-by-sex interaction | 0.119 | 0.000 | 0.888 | 0.888 |
| Hippocampal tail | L | Diagnostic group | 2.123 | 0.003 | 0.120 | 0.156 |
|  | L | Sex | 4.117 | 0.003 | **0.043** | 0.068 |
|  | L | Group-by-sex interaction | 1.731 | 0.002 | 0.178 | 0.220 |
|  | R | Diagnostic group | 0.889 | 0.001 | 0.411 | 0.446 |
|  | R | Sex | 13.669 | 0.009 | **2.26e-04** | **0.002** |
|  | R | Group-by-sex interaction | 2.337 | 0.003 | 0.097 | 0.130 |
| Molecular layer | L | Diagnostic group | 5.846 | 0.008 | **0.003** | **0.011** |
|  | L | Sex | 9.714 | 0.006 | **0.002** | **0.009** |
|  | L | Group-by-sex interaction | 4.594 | 0.006 | **0.010** | **0.024** |
|  | R | Diagnostic group | 8.500 | 0.011 | **2.14e-04** | **0.002** |
|  | R | Sex | 5.649 | 0.004 | **0.018** | **0.034** |
|  | R | Group-by-sex interaction | 3.753 | 0.005 | **0.024** | **0.044** |
| Parasubiculum | L | Diagnostic group | 1.083 | 0.001 | 0.339 | 0.372 |
|  | L | Sex | 6.549 | 0.004 | **0.011** | **0.024** |
|  | L | Group-by-sex interaction | 1.147 | 0.002 | 0.318 | 0.354 |
|  | R | Diagnostic group | 0.163 | 0.000 | 0.849 | 0.868 |
|  | R | Sex | 11.783 | 0.008 | **0.001** | **0.003** |
|  | R | Group-by-sex interaction | 4.200 | 0.006 | **0.015** | **0.032** |
| Presubiculum | L | Diagnostic group | 1.479 | 0.002 | 0.228 | 0.270 |
|  | L | Sex | 13.299 | 0.009 | **2.75e-04** | **0.002** |
|  | L | Group-by-sex interaction | 2.574 | 0.003 | 0.077 | 0.105 |
|  | R | Diagnostic group | 2.907 | 0.004 | 0.055 | 0.082 |
|  | R | Sex | 14.876 | 0.010 | **1.20e-04** | **0.001** |
|  | R | Group-by-sex interaction | 4.648 | 0.006 | **0.010** | **0.024** |
| Subiculum | L | Diagnostic group | 0.724 | 0.001 | 0.485 | 0.518 |
|  | L | Sex | 4.536 | 0.003 | **0.033** | 0.057 |
|  | L | Group-by-sex interaction | 1.546 | 0.002 | 0.213 | 0.256 |
|  | R | Diagnostic group | 1.392 | 0.002 | 0.249 | 0.286 |
|  | R | Sex | 7.415 | 0.005 | **0.007** | **0.018** |
|  | R | Group-by-sex interaction | 4.786 | 0.006 | **0.008** | **0.023** |
| Hippocampus | L | Diagnostic group | 5.775 | 0.008 | **0.003** | **0.011** |
|  | L | Sex | 17.738 | 0.012 | **2.69e-05** | **4.19e-04** |
|  | L | Group-by-sex interaction | 5.330 | 0.007 | **0.005** | **0.016** |
|  | R | Diagnostic group | 6.167 | 0.008 | **0.002** | **0.009** |
|  | R | Sex | 16.489 | 0.011 | **5.15e-05** | **0.001** |
|  | R | Group-by-sex interaction | 5.663 | 0.007 | **0.004** | **0.012** |

The statistical results are based on analysis of covariance (adjusted for age, age^2^ and intracranial volume), Abbreviation: CA = cornu ammonis, GC-ML-DG = granule cells in the molecular layer of the dentate gyrus, HATA = hippocampal-amygdaloid transition area, FDR = false discovery rate. Significant results are highlighted in bold.

| **Subfields** | **Side** | **CTL** | **CI** | **BD** | **CI** | **SZ** | **CI** | **F-value** | **partial η^2^** | **p-**  **value** | **p_FDR_** |
| --- | --- | --- | --- | --- | --- | --- | --- | --- | --- | --- | --- |
| CA1 | L | 675.53 ± 2.39 | [670.85, 680.21] | 654.82 ± 3.68 | [647.61, 662.04] | 647.20 ± 3.10 | [641.12, 653.27] | 28.91 | 0.037 | **4.78e-13** | **1.78e-12** |
|  | R | 710.78 ± 2.53 | [705.81, 715.75] | 688.35 ± 3.91 | [680.69, 696.01] | 680.09 ± 3.29 | [673.63, 686.54] | 30.09 | 0.038 | **1.53e-13** | **6.64e-13** |
| CA3 | L | 212.09 ± 0.98 | [210.17, 214.00] | 207.28 ± 1.50 | [204.33, 210.23] | 205.88 ± 1.27 | [203.39, 208.36] | 8.53 | 0.011 | **2.06e-04** | **2.82e-04** |
|  | R | 230.77 ± 1.03 | [228.76, 232.79] | 225.18 ± 1.59 | [222.07, 228.30] | 222.57 ± 1.34 | [219.95, 225.20] | 12.69 | 0.016 | **3.43e-06** | **5.57e-06** |
| CA4 | L | 254.05 ± 0.82 | [252.45, 255.66] | 248.23 ± 1.26 | [245.76, 250.70] | 245.17 ± 1.06 | [243.09, 247.25] | 23.33 | 0.030 | **1.05e-10** | **2.48e-10** |
|  | R | 262.49 ± 0.82 | [260.89, 264.10] | 256.63 ± 1.26 | [254.15, 259.11] | 252.95 ± 1.06 | [250.86, 255.04] | 26.28 | 0.034 | **6.04e-12** | **1.57e-11** |
| Fimbria | L | 90.49 ± 0.64 | [89.22, 91.75] | 87.75 ± 0.99 | [85.80, 89.70] | 87.48 ± 0.84 | [85.84, 89.12] | 5.03 | 0.007 | **0.007** | **0.008** |
|  | R | 85.86 ± 0.62 | [84.65, 87.08] | 84.79 ± 0.96 | [82.92, 86.67] | 83.46 ± 0.80 | [81.88, 85.04] | 2.78 | 0.004 | 0.062 | 0.065 |
| GC-ML-DG | L | 299.43 ± 0.94 | [297.59, 301.26] | 291.58 ± 1.45 | [288.75, 294.42] | 288.39 ± 1.22 | [286.00, 290.78] | 28.12 | 0.036 | **1.02e-12** | **3.32e-12** |
|  | R | 308.49 ± 0.94 | [306.65, 310.33] | 300.74 ± 1.45 | [297.91, 303.58] | 296.35 ± 1.22 | [293.96, 298.74] | 32.75 | 0.041 | **1.19e-14** | **7.75e-14** |
| HATA | L | 66.42 ± 0.33 | [65.78, 67.06] | 63.76 ± 0.50 | [62.78, 64.75] | 62.64 ± 0.42 | [61.82, 63.47] | 27.18 | 0.035 | **2.53e-12** | **7.31e-12** |
|  | R | 69.38 ± 0.34 | [68.71, 70.06] | 67.20 ± 0.53 | [66.16, 68.24] | 66.02 ± 0.45 | [65.15, 66.90] | 18.88 | 0.024 | **7.97e-09** | **1.59e-08** |
| Hippocampal tail | L | 587.55 ± 2.62 | [582.41, 592.69] | 576.09 ± 4.04 | [568.16, 584.02] | 565.31 ± 3.40 | [558.64, 571.99] | 13.51 | 0.018 | **1.52e-06** | **2.64e-06** |
|  | R | 596.10 ± 2.61 | [590.99, 601.21] | 590.66 ± 4.02 | [582.78, 598.55] | 577.21 ± 3.38 | [570.57, 583.85] | 9.77 | 0.013 | **6.09e-05** | **8.80e-05** |
| Hippocampal fissure | L | 145.51 ± 0.86 | [143.83, 147.20] | 146.90 ± 1.32 | [144.31, 149.49] | 148.77 ± 1.11 | [146.58, 150.95] | 2.66 | 0.004 | 0.070 | 0.070 |
|  | R | 151.06 ± 0.86 | [149.38, 152.74] | 154.54 ± 1.32 | [151.94, 157.13] | 154.43 ± 1.11 | [152.25, 156.62] | 3.95 | 0.005 | **0.020** | **0.021** |
| Molecular layer | L | 585.27 ± 1.70 | [581.94, 588.60] | 570.41 ± 1.32 | [565.28, 575.55] | 564.63 ± 2.21 | [560.30, 568.95] | 30.09 | 0.038 | **1.53e-13** | **6.64e-13** |
|  | R | 600.31 ± 1.73 | [596.92, 603.71] | 583.47 ± 2.67 | [578.23, 588.71] | 575.60 ± 2.25 | [571.19, 580.02] | 40.64 | 0.051 | **6.44e-18** | **1.68e-16** |
| Parasubiculum | L | 70.34 ± 0.43 | [69.49, 71.19] | 69.00 ± 0.67 | [67.69, 70.31] | 68.06 ± 0.56 | [66.96, 69.16] | 5.32 | 0.007 | **0.005** | **0.006** |
|  | R | 67.52 ± 0.41 | [66.72, 68.32] | 66.17 ± 0.63 | [64.93, 67.40] | 64.79 ± 0.53 | [63.75, 65.83] | 8.36 | 0.011 | **2.45e-04** | **3.19e-04** |
| Presubiculum | L | 331.44 ± 1.35 | [328.80, 334.08] | 324.15 ± 2.08 | [320.08, 328.22] | 320.75 ± 1.75 | [317.32, 324.18] | 12.61 | 0.016 | **3.724-06** | **5.68e-06** |
|  | R | 308.90 ± 1.17 | [306.59, 311.20] | 302.24 ± 1.81 | [298.69, 305.79] | 296.15 ± 1.53 | [293.15, 299.14] | 22.16 | 0.028 | **3.254-10** | **7.05e-10** |
| Subiculum | L | 454.25 ± 1.59 | [451.12, 457.37] | 448.40 ± 2.46 | [443.58, 453.22] | 445.01 ± 2.07 | [440.96, 449.07] | 6.57 | 0.009 | **0.002** | **0.002** |
|  | R | 445.43 ± 1.43 | [442.63, 448.24] | 438.83 ± 2.21 | [434.50, 443.16] | 432.39 ± 1.86 | [428.75, 436.03] | 15.57 | 0.020 | **2.03e-07** | **3.76e-07** |
| Hippocampus | L | 3626.95 ± 9.66 | [3608.00, 3645.90] | 3541.58 ± 14.90 | [3512.35, 3570.80] | 3500.46 ± 12.55 | [3475.84, 3525.07] | 34.11 | 0.043 | **3.23e-15** | **2.80e-14** |
|  | R | 3686.06 ± 9.78 | [3666.87, 3705.25] | 3604.22 ± 15.08 | [3574.63, 3633.80] | 3547.60 ± 12.70 | [3522.68, 3572.52] | 38.47 | 0.048 | **5.05e-17** | **6.57e-16** |

**Table S2| Diagnostic group effects on total hippocampal and subfield volumes assessed using analysis of covariance.**

The statistical results are based on analysis of covariance (adjusted for age, age^2^ and intracranial volume), Abbreviation: L = left, R = right, CTL = healthy controls, BD = bipolar disorders, SZ = schizophrenia spectrum disorders, CA = cornu ammonis, GC-ML-DG = granule cells in the molecular layer of the dentate gyrus, HATA = hippocampal-amygdaloid transition area, FDR = false discovery rate. Significant results are highlighted in bold.

**Table S3| Sex, diagnostic group and sex-by-diagnostic group effects on total hippocampal and subfield volumes assessed using analysis of covariance, without adjustment for intracranial volume.**

| **Subfield** | **Variable** | **F-value** | **partial η ^2^** | **p-value** | **p_FDR_-value** |
| --- | --- | --- | --- | --- | --- |
| CA1 | Diagnostic group | 9.813 | 0.013 | **5.83e-05** | **1.62e-04** |
|  | Sex | 184.261 | 0.109 | **1.08e-39** | **1.41e-38** |
|  | Group-by-sex interaction | 1.633 | 0.002 | 0.196 | 0.218 |
| CA3 | Diagnostic group | 3.667 | 0.005 | **0.026** | **0.044** |
|  | Sex | 107.390 | 0.066 | **2.34e-24** | **9.11e-24** |
|  | Group-by-sex interaction | 2.435 | 0.003 | 0.088 | 0.107 |
| CA4 | Diagnostic group | 6.447 | 0.008 | **0.002** | **0.003** |
|  | Sex | 148.217 | 0.089 | **1.35e-32** | **6.57e-32** |
|  | Group-by-sex interaction | 3.373 | 0.004 | **0.035** | 0.056 |
| Fimbria | Diagnostic group | 0.291 | 3.85e-04 | 0.747 | 0.767 |
|  | Sex | 161.307 | 0.096 | **3.42e-35** | **2.22e-34** |
|  | Group-by-sex interaction | 3.150 | 0.004 | **0.043** | 0.065 |
| GC-ML-DG | Diagnostic group | 7.525 | 0.010 | **0.001** | **0.001** |
|  | Sex | 171.779 | 0.102 | **2.98e-37** | **2.90e-36** |
|  | Group-by-sex interaction | 3.872 | 0.005 | **0.021** | **0.039** |
| HATA | Diagnostic group | 9.533 | 0.012 | **7.69e-05** | **2.00e-04** |
|  | Sex | 141.521 | 0.086 | **2.92e-31** | **1.27e-30** |
|  | Group-by-sex interaction | 3.259 | 0.004 | **0.039** | 0.060 |
| Hippocampal tail | Diagnostic group | 2.952 | 0.004 | 0.053 | 0.071 |
|  | Sex | 89.282 | 0.056 | **1.25e-20** | **4.07e-20** |
|  | Group-by-sex interaction | 1.980 | 0.003 | 0.138 | 0.159 |
| Hippocampal fissure | Diagnostic group | 1.486 | 0.002 | 0.227 | 0.246 |
|  | Sex | 85.794 | 0.054 | **6.63e-20** | **1.99e-19** |
|  | Group-by-sex interaction | 0.184 | 2.43e-04 | 0.832 | 0.832 |
| Molecular layer | Diagnostic group | 9.011 | 0.012 | **1.29e-04** | **3.14e-04** |
|  | Sex | 185.684 | 0.109 | **5.73e-40** | **1.12e-38** |
|  | Group-by-sex interaction | 3.066 | 0.004 | **0.047** | 0.065 |
| Parasubiculum | Diagnostic group | 1.353 | 0.002 | 0.259 | 0.273 |
|  | Sex | 97.613 | 0.061 | **2.38e-22** | **8.42e-22** |
|  | Group-by-sex interaction | 2.527 | 0.003 | 0.080 | 0.104 |
| Presubiculum | Diagnostic group | 4.002 | 0.005 | **0.018** | **0.036** |
|  | Sex | 166.675 | 0.099 | **3.00e-36** | **2.34e-35** |
|  | Group-by-sex interaction | 3.109 | 0.004 | **0.045** | 0.065 |
| Subiculum | Diagnostic group | 2.497 | 0.003 | 0.083 | 0.104 |
|  | Sex | 152.782 | 0.092 | **1.67e-33** | **9.29e-33** |
|  | Group-by-sex interaction | 2.282 | 0.003 | 0.102 | 0.121 |
| Hippocampus | Diagnostic group | 7.854 | 0.010 | **4.04e-04** | **0.001** |
|  | Sex | 232.614 | 0.133 | **5.78e-49** | **2.26e-47** |
|  | Group-by-sex interaction | 3.830 | 0.005 | **0.022** | **0.039** |

The statistical results are based on analysis of covariance (adjusted for age and age^2^) Abbreviation: CA = cornu ammonis, GC-ML-DG = granule cells in the molecular layer of the dentate gyrus, HATA = hippocampal-amygdaloid transition area, FDR = false discovery rate. Significant results are highlighted in bold.

**Table S4| Diagnostic group and sex differences in hippocampal volumes.**

| **Subfields** | **Contrast*** | **Diagnostic group** | **Estimate** | **S.E.** | **t-value** | **p-value** |
| --- | --- | --- | --- | --- | --- | --- |
| CA1 | Female vs Female | CTL - BD | 36.171 | 11.259 | 3.213 | **0.017** |
|  | Female vs Female | CTL - SZ | 40.654 | 11.303 | 3.597 | **0.004** |
|  | Female vs Female | BD - SZ | 4.482 | 12.753 | 0.351 | 0.999 |
|  | Female vs Male | CTL - CTL | -40.310 | 10.042 | -4.014 | **0.001** |
|  | Female vs Male | CTL - BD | 8.544 | 13.510 | 0.632 | 0.989 |
|  | Female vs Male | CTL - SZ | 32.217 | 10.855 | 2.968 | **0.036** |
|  | Female vs Male | BD - CTL | -76.481 | 11.766 | -6.500 | **1.63e-09** |
|  | Female vs Male | BD - BD | -27.628 | 14.730 | -1.876 | 0.418 |
|  | Female vs Male | BD - SZ | -3.954 | 12.373 | -0.320 | 1.000 |
|  | Female vs Male | SZ - CTL | -80.964 | 12.056 | -6.715 | **3.97e-10** |
|  | Female vs Male | SZ - BD | -32.110 | 14.967 | -2.145 | 0.265 |
|  | Female vs Male | SZ - SZ | -8.437 | 12.571 | -0.671 | 0.985 |
|  | Male vs Male | CTL - BD | 48.853 | 12.430 | 3.930 | **0.001** |
|  | Male vs Male | CTL - SZ | 72.527 | 9.715 | 7.465 | **2.15e-12** |
|  | Male vs Male | BD - SZ | 23.673 | 13.228 | 1.790 | 0.473 |
| CA3 | Female vs Female | CTL - BD | 10.614 | 4.445 | 2.388 | 0.161 |
|  | Female vs Female | CTL - SZ | 6.775 | 4.463 | 1.518 | 0.653 |
|  | Female vs Female | BD - SZ | -3.840 | 5.035 | -0.763 | 0.974 |
|  | Female vs Male | CTL - CTL | -7.630 | 3.965 | -1.925 | 0.387 |
|  | Female vs Male | CTL - BD | 1.146 | 5.334 | 0.215 | 1.000 |
|  | Female vs Male | CTL - SZ | 12.304 | 4.286 | 2.871 | **0.048** |
|  | Female vs Male | BD - CTL | -18.245 | 4.645 | -3.928 | **0.001** |
|  | Female vs Male | BD - BD | -9.468 | 5.816 | -1.628 | 0.580 |
|  | Female vs Male | BD - SZ | 1.689 | 4.885 | 0.346 | 0.999 |
|  | Female vs Male | SZ - CTL | -14.405 | 4.760 | -3.026 | **0.030** |
|  | Female vs Male | SZ - BD | -5.628 | 5.909 | -0.952 | 0.933 |
|  | Female vs Male | SZ - SZ | 5.529 | 4.963 | 1.114 | 0.876 |
|  | Male vs Male | CTL - BD | 8.777 | 4.908 | 1.788 | 0.474 |
|  | Male vs Male | CTL - SZ | 19.934 | 3.836 | 5.197 | **3.42e-06** |
|  | Male vs Male | BD - SZ | 11.157 | 5.223 | 2.136 | 0.269 |
| CA4 | Female vs Female | CTL - BD | 9.970 | 3.693 | 2.700 | 0.076 |
|  | Female vs Female | CTL - SZ | 9.783 | 3.707 | 2.639 | 0.089 |
|  | Female vs Female | BD - SZ | -0.187 | 4.183 | -0.045 | 1.000 |
|  | Female vs Male | CTL - CTL | -6.465 | 3.293 | -1.963 | 0.364 |
|  | Female vs Male | CTL - BD | 5.912 | 4.431 | 1.334 | 0.766 |
|  | Female vs Male | CTL - SZ | 18.260 | 3.560 | 5.129 | **4.88e-06** |
|  | Female vs Male | BD - CTL | -16.434 | 3.859 | -4.259 | **3.14e-04** |
|  | Female vs Male | BD - BD | -4.057 | 4.831 | -0.840 | 0.960 |
|  | Female vs Male | BD - SZ | 8.291 | 4.058 | 2.043 | 0.318 |
|  | Female vs Male | SZ - CTL | -16.247 | 3.954 | -4.109 | **0.001** |
|  | Female vs Male | SZ - BD | -3.870 | 4.909 | -0.788 | 0.970 |
|  | Female vs Male | SZ - SZ | 8.478 | 4.123 | 2.056 | 0.311 |
|  | Male vs Male | CTL - BD | 12.377 | 4.077 | 3.036 | **0.029** |
|  | Male vs Male | CTL - SZ | 24.725 | 3.186 | 7.760 | **2.94e-13** |
|  | Male vs Male | BD - SZ | 12.348 | 4.338 | 2.846 | 0.051 |
| Fimbria | Female vs Female | CTL - BD | 0.970 | 2.794 | 0.347 | 0.999 |
|  | Female vs Female | CTL - SZ | -0.413 | 2.805 | -0.147 | 1.000 |
|  | Female vs Female | BD - SZ | -1.383 | 3.165 | -0.437 | 0.998 |
|  | Female vs Male | CTL - CTL | -11.953 | 2.492 | -4.797 | **2.61e-05** |
|  | Female vs Male | CTL - BD | -5.532 | 3.352 | -1.650 | 0.565 |
|  | Female vs Male | CTL - SZ | -2.247 | 2.694 | -0.834 | 0.961 |
|  | Female vs Male | BD - CTL | -12.923 | 2.920 | -4.426 | **1.49e-04** |
|  | Female vs Male | BD - BD | -6.503 | 3.655 | -1.779 | 0.480 |
|  | Female vs Male | BD - SZ | -3.217 | 3.070 | -1.048 | 0.902 |
|  | Female vs Male | SZ - CTL | -11.540 | 2.992 | -3.857 | **0.002** |
|  | Female vs Male | SZ - BD | -5.119 | 3.714 | -1.378 | 0.740 |
|  | Female vs Male | SZ - SZ | -1.834 | 3.119 | -0.588 | 0.992 |
|  | Male vs Male | CTL - BD | 6.420 | 3.084 | 2.081 | 0.297 |
|  | Male vs Male | CTL - SZ | 9.706 | 2.411 | 4.026 | **0.001** |
|  | Male vs Male | BD - SZ | 3.286 | 3.282 | 1.001 | 0.918 |
| GC-ML-DG | Female vs Female | CTL - BD | 13.280 | 4.249 | 3.126 | **0.022** |
|  | Female vs Female | CTL - SZ | 12.320 | 4.265 | 2.888 | **0.045** |
|  | Female vs Female | BD - SZ | -0.960 | 4.812 | -0.199 | 1.000 |
|  | Female vs Male | CTL - CTL | -9.628 | 3.789 | -2.541 | 0.113 |
|  | Female vs Male | CTL - BD | 7.009 | 5.098 | 1.375 | 0.742 |
|  | Female vs Male | CTL - SZ | 21.475 | 4.096 | 5.243 | **2.69e-06** |
|  | Female vs Male | BD - CTL | -22.907 | 4.440 | -5.160 | **4.17e-06** |
|  | Female vs Male | BD - BD | -6.271 | 5.559 | -1.128 | 0.870 |
|  | Female vs Male | BD - SZ | 8.195 | 4.669 | 1.755 | 0.495 |
|  | Female vs Male | SZ - CTL | -21.948 | 4.550 | -4.824 | **2.28e-05** |
|  | Female vs Male | SZ - BD | -5.311 | 5.648 | -0.940 | 0.936 |
|  | Female vs Male | SZ - SZ | 9.155 | 4.744 | 1.930 | 0.384 |
|  | Male vs Male | CTL - BD | 16.637 | 4.691 | 3.547 | **0.005** |
|  | Male vs Male | CTL - SZ | 31.102 | 3.666 | 8.484 | **7.07e-14** |
|  | Male vs Male | BD - SZ | 14.465 | 4.992 | 2.898 | **0.044** |
| HATA | Female vs Female | CTL - BD | 5.662 | 1.494 | 3.789 | **0.002** |
|  | Female vs Female | CTL - SZ | 4.677 | 1.500 | 3.118 | **0.023** |
|  | Female vs Female | BD - SZ | -0.984 | 1.693 | -0.582 | 0.992 |
|  | Female vs Male | CTL - CTL | -5.202 | 1.333 | -3.903 | **0.001** |
|  | Female vs Male | CTL - BD | -1.845 | 1.793 | -1.029 | 0.908 |
|  | Female vs Male | CTL - SZ | 3.689 | 1.441 | 2.561 | 0.108 |
|  | Female vs Male | BD - CTL | -10.864 | 1.562 | -6.957 | **7.74e-11** |
|  | Female vs Male | BD - BD | -7.506 | 1.955 | -3.839 | **0.002** |
|  | Female vs Male | BD - SZ | -1.972 | 1.642 | -1.201 | 0.837 |
|  | Female vs Male | SZ - CTL | -9.880 | 1.600 | -6.174 | **1.28e-08** |
|  | Female vs Male | SZ - BD | -6.522 | 1.987 | -3.283 | **0.013** |
|  | Female vs Male | SZ - SZ | -0.988 | 1.668 | -0.592 | 0.992 |
|  | Male vs Male | CTL - BD | 3.358 | 1.650 | 2.035 | 0.323 |
|  | Male vs Male | CTL - SZ | 8.892 | 1.289 | 6.896 | **1.18e-10** |
|  | Male vs Male | BD - SZ | 5.534 | 1.756 | 3.152 | **0.020** |
| Fissure | Female vs Female | CTL - BD | -6.078 | 3.795 | -1.602 | 0.598 |
|  | Female vs Female | CTL - SZ | -6.148 | 3.810 | -1.614 | 0.590 |
|  | Female vs Female | BD - SZ | -0.069 | 4.299 | -0.016 | 1.000 |
|  | Female vs Male | CTL - CTL | -17.950 | 3.385 | -5.303 | **1.95e-06** |
|  | Female vs Male | CTL - BD | -21.170 | 4.554 | -4.649 | **5.32e-05** |
|  | Female vs Male | CTL - SZ | -24.889 | 3.659 | -6.803 | **2.21e-10** |
|  | Female vs Male | BD - CTL | -11.872 | 3.966 | -2.994 | **0.033** |
|  | Female vs Male | BD - BD | -15.091 | 4.965 | -3.039 | **0.029** |
|  | Female vs Male | BD - SZ | -18.811 | 4.171 | -4.510 | **1.02e-04** |
|  | Female vs Male | SZ - CTL | -11.802 | 4.064 | -2.904 | **0.043** |
|  | Female vs Male | SZ - BD | -15.022 | 5.045 | -2.978 | **0.035** |
|  | Female vs Male | SZ - SZ | -18.741 | 4.237 | -4.423 | **1.51e-04** |
|  | Male vs Male | CTL - BD | -3.219 | 4.190 | -0.768 | 0.973 |
|  | Male vs Male | CTL - SZ | -6.939 | 3.275 | -2.119 | 0.278 |
|  | Male vs Male | BD - SZ | -3.720 | 4.459 | -0.834 | 0.961 |
| Tail | Female vs Female | CTL - BD | 7.242 | 12.172 | 0.595 | 0.991 |
|  | Female vs Female | CTL - SZ | 21.742 | 12.219 | 1.779 | 0.479 |
|  | Female vs Female | BD - SZ | 14.500 | 13.786 | 1.052 | 0.900 |
|  | Female vs Male | CTL - CTL | -33.201 | 10.856 | -3.058 | **0.027** |
|  | Female vs Male | CTL - BD | -7.323 | 14.605 | -0.501 | 0.996 |
|  | Female vs Male | CTL - SZ | 22.242 | 11.734 | 1.895 | 0.405 |
|  | Female vs Male | BD - CTL | -40.443 | 12.719 | -3.180 | **0.019** |
|  | Female vs Male | BD - BD | -14.565 | 15.924 | -0.915 | 0.943 |
|  | Female vs Male | BD - SZ | 15.000 | 13.376 | 1.121 | 0.873 |
|  | Female vs Male | SZ - CTL | -54.943 | 13.034 | -4.216 | **3.79e-04** |
|  | Female vs Male | SZ - BD | -29.065 | 16.180 | -1.796 | 0.468 |
|  | Female vs Male | SZ - SZ | 0.500 | 13.590 | 0.037 | 1.000 |
|  | Male vs Male | CTL - BD | 25.878 | 13.437 | 1.926 | 0.387 |
|  | Male vs Male | CTL - SZ | 55.443 | 10.502 | 5.279 | **2.21e-06** |
|  | Male vs Male | BD - SZ | 29.565 | 14.300 | 2.067 | 0.305 |
| Molecular layer | Female vs Female | CTL - BD | 26.028 | 7.943 | 3.277 | **0.014** |
|  | Female vs Female | CTL - SZ | 26.871 | 7.974 | 3.370 | **0.010** |
|  | Female vs Female | BD - SZ | 0.843 | 8.997 | 0.094 | 1.000 |
|  | Female vs Male | CTL - CTL | -20.869 | 7.084 | -2.946 | **0.038** |
|  | Female vs Male | CTL - BD | 14.861 | 9.531 | 1.559 | 0.626 |
|  | Female vs Male | CTL - SZ | 38.029 | 7.658 | 4.966 | **1.13e-05** |
|  | Female vs Male | BD - CTL | -46.897 | 8.300 | -5.650 | **2.86e-07** |
|  | Female vs Male | BD - BD | -11.167 | 10.392 | -1.075 | 0.892 |
|  | Female vs Male | BD - SZ | 12.001 | 8.729 | 1.375 | 0.742 |
|  | Female vs Male | SZ - CTL | -47.740 | 8.505 | -5.613 | **3.53e-07** |
|  | Female vs Male | SZ - BD | -12.010 | 10.559 | -1.137 | 0.866 |
|  | Female vs Male | SZ - SZ | 11.158 | 8.868 | 1.258 | 0.808 |
|  | Male vs Male | CTL - BD | 35.730 | 8.769 | 4.075 | **0.001** |
|  | Male vs Male | CTL - SZ | 58.898 | 6.854 | 8.594 | **6.62e-14** |
|  | Male vs Male | BD - SZ | 23.168 | 9.332 | 2.483 | 0.130 |
| Parasubiculum | Female vs Female | CTL - BD | 1.855 | 1.826 | 1.016 | 0.913 |
|  | Female vs Female | CTL - SZ | 1.697 | 1.833 | 0.926 | 0.940 |
|  | Female vs Female | BD - SZ | -0.158 | 2.068 | -0.077 | 1.000 |
|  | Female vs Male | CTL - CTL | -5.568 | 1.628 | -3.419 | **0.008** |
|  | Female vs Male | CTL - BD | -2.392 | 2.191 | -1.092 | 0.885 |
|  | Female vs Male | CTL - SZ | 1.866 | 1.760 | 1.060 | 0.897 |
|  | Female vs Male | BD - CTL | -7.423 | 1.908 | -3.891 | **0.001** |
|  | Female vs Male | BD - BD | -4.247 | 2.389 | -1.778 | 0.480 |
|  | Female vs Male | BD - SZ | 0.010 | 2.006 | 0.005 | 1.000 |
|  | Female vs Male | SZ - CTL | -7.265 | 1.955 | -3.716 | **0.003** |
|  | Female vs Male | SZ - BD | -4.088 | 2.427 | -1.685 | 0.542 |
|  | Female vs Male | SZ - SZ | 0.169 | 2.038 | 0.083 | 1.000 |
|  | Male vs Male | CTL - BD | 3.176 | 2.016 | 1.576 | 0.615 |
|  | Male vs Male | CTL - SZ | 7.433 | 1.575 | 4.719 | **3.82e-05** |
|  | Male vs Male | BD - SZ | 4.257 | 2.145 | 1.985 | 0.352 |
| Presubiculum | Female vs Female | CTL - BD | 9.863 | 5.657 | 1.744 | 0.503 |
|  | Female vs Female | CTL - SZ | 11.215 | 5.679 | 1.975 | 0.357 |
|  | Female vs Female | BD - SZ | 1.352 | 6.407 | 0.211 | 1.000 |
|  | Female vs Male | CTL - CTL | -20.939 | 5.045 | -4.150 | **0.001** |
|  | Female vs Male | CTL - BD | -3.880 | 6.788 | -0.572 | 0.993 |
|  | Female vs Male | CTL - SZ | 11.482 | 5.454 | 2.105 | 0.285 |
|  | Female vs Male | BD - CTL | -30.802 | 5.911 | -5.211 | **3.19e-06** |
|  | Female vs Male | BD - BD | -13.743 | 7.401 | -1.857 | 0.429 |
|  | Female vs Male | BD - SZ | 1.619 | 6.217 | 0.260 | 1.000 |
|  | Female vs Male | SZ - CTL | -32.153 | 6.057 | -5.308 | 0.000 |
|  | Female vs Male | SZ - BD | -15.095 | 7.520 | -2.007 | 0.338 |
|  | Female vs Male | SZ - SZ | 0.267 | 6.316 | 0.042 | 1.000 |
|  | Male vs Male | CTL - BD | 17.059 | 6.245 | 2.731 | 0.070 |
|  | Male vs Male | CTL - SZ | 32.421 | 4.881 | 6.642 | **6.45e-10** |
|  | Male vs Male | BD - SZ | 15.362 | 6.646 | 2.311 | 0.190 |
| Subiculum | Female vs Female | CTL - BD | 8.872 | 6.999 | 1.268 | 0.803 |
|  | Female vs Female | CTL - SZ | 8.820 | 7.027 | 1.255 | 0.809 |
|  | Female vs Female | BD - SZ | -0.053 | 7.928 | -0.007 | 1.000 |
|  | Female vs Male | CTL - CTL | -16.168 | 6.243 | -2.590 | 0.100 |
|  | Female vs Male | CTL - BD | -1.487 | 8.399 | -0.177 | 1.000 |
|  | Female vs Male | CTL - SZ | 15.959 | 6.748 | 2.365 | 0.169 |
|  | Female vs Male | BD - CTL | -25.040 | 7.314 | -3.423 | **0.008** |
|  | Female vs Male | BD - BD | -10.359 | 9.157 | -1.131 | 0.868 |
|  | Female vs Male | BD - SZ | 7.086 | 7.692 | 0.921 | 0.941 |
|  | Female vs Male | SZ - CTL | -24.988 | 7.495 | -3.334 | **0.011** |
|  | Female vs Male | SZ - BD | -10.307 | 9.304 | -1.108 | 0.878 |
|  | Female vs Male | SZ - SZ | 7.139 | 7.815 | 0.913 | 0.943 |
|  | Male vs Male | CTL - BD | 14.681 | 7.727 | 1.900 | 0.403 |
|  | Male vs Male | CTL - SZ | 32.126 | 6.040 | 5.319 | **1.78e-06** |
|  | Male vs Male | BD - SZ | 17.446 | 8.223 | 2.121 | 0.277 |
| Hippocampus | Female vs Female | CTL - BD | 130.523 | 45.797 | 2.850 | *0.050* |
|  | Female vs Female | CTL - SZ | 144.056 | 45.975 | 3.133 | **0.022** |
|  | Female vs Female | BD - SZ | 13.533 | 51.873 | 0.261 | 1.000 |
|  | Female vs Male | CTL - CTL | -178.165 | 40.845 | -4.362 | **1.99e-04** |
|  | Female vs Male | CTL - BD | 14.916 | 54.953 | 0.271 | 1.000 |
|  | Female vs Male | CTL - SZ | 175.372 | 44.152 | 3.972 | **0.001** |
|  | Female vs Male | BD - CTL | -308.688 | 47.857 | -6.450 | **2.25e-09** |
|  | Female vs Male | BD - BD | -115.607 | 59.916 | -1.929 | 0.384 |
|  | Female vs Male | BD - SZ | 44.849 | 50.330 | 0.891 | 0.949 |
|  | Female vs Male | SZ - CTL | -322.221 | 49.040 | -6.571 | **1.03e-09** |
|  | Female vs Male | SZ - BD | -129.140 | 60.878 | -2.121 | 0.277 |
|  | Female vs Male | SZ - SZ | 31.316 | 51.132 | 0.612 | 0.990 |
|  | Male vs Male | CTL - BD | 193.081 | 50.560 | 3.819 | **0.002** |
|  | Male vs Male | CTL - SZ | 353.537 | 39.516 | 8.947 | **6.36e-14** |
|  | Male vs Male | BD - SZ | 160.456 | 53.805 | 2.982 | **0.034** |

*Post hoc tukey tests were performed to contrast volume differences between and within the sexes across diagnostic groups. Statistical results are based on analysis of covariance (adjusted for age, age^2^, intracranial volume). P value adjustment is based om the tukey method for comparing a family of 6 estimates. Abbreviation: CTL = healthy controls, BD = bipolar disorders, SZ = schizophrenia spectrum disorders, CA = cornu ammonis, GC-ML-DG = granule cells in the molecular layer of the dentate gyrus, HATA = hippocampal-amygdaloid transition area, S.E. = standard error. Significant results are highlighted in bold.

**Table S5| Sex-by-diagnostic subgroup differences in hippocampal volumes.**

| **Subfield** | **Group** | **Estimate** | **S.E.** | **t-value** | **p-value** | **p_FDR_-value** | **Cohen *d* [95% CI]** |
| --- | --- | --- | --- | --- | --- | --- | --- |
| CA1 | BP I | -17.891 | 20.261 | -0.883 | 0.377 | 0.721 | -0.046 [-0.149,0.056] |
|  | BP II | -5.325 | 25.852 | -0.206 | 0.837 | 0.942 | -0.011 [-0.113,0.092] |
|  | OTP | -1.767 | 24.774 | -0.071 | 0.943 | 0.966 | -0.004 [-0.106,0.099] |
|  | SCZ-AF | -40.874 | 34.74 | -1.177 | 0.240 | 0.599 | -0.062 [-0.164,0.041] |
|  | SCZ | -46.719 | 18.999 | -2.459 | **0.014** | 0.152 | -0.129 [-0.231,-0.026] |
| CA3 | BP I | 3.78 | 7.994 | 0.473 | 0.636 | 0.889 | 0.025 [-0.078,0.127] |
|  | BP II | 5.649 | 10.2 | 0.554 | 0.580 | 0.857 | 0.029 [-0.074,0.132] |
|  | OTP | -2.734 | 9.775 | -0.28 | 0.780 | 0.942 | -0.015 [-0.117,0.088] |
|  | SCZ-AF | -26.31 | 13.707 | -1.919 | 0.055 | 0.235 | -0.100 [-0.203,0.002] |
|  | SCZ | -15.779 | 7.496 | -2.105 | **0.035** | 0.235 | -0.110 [-0.213,-0.007] |
| CA4 | BP I | -3.35 | 6.636 | -0.505 | 0.614 | 0.886 | -0.026 [-0.129,0.076] |
|  | BP II | 3.137 | 8.468 | 0.37 | 0.711 | 0.942 | 0.019 [-0.083,0.122] |
|  | OTP | -7.841 | 8.115 | -0.966 | 0.334 | 0.679 | -0.051 [-0.153,0.052] |
|  | SCZ-AF | -22.119 | 11.379 | -1.944 | 0.052 | 0.235 | -0.102 [-0.204,0.001] |
|  | SCZ | -18.051 | 6.223 | -2.901 | **0.004** | **0.049** | -0.152 [-0.255,-0.049] |
| Fimbria | BP I | -9.351 | 4.994 | -1.873 | 0.061 | 0.235 | -0.098 [-0.201,0.005] |
|  | BP II | -2.11 | 6.372 | -0.331 | 0.741 | 0.942 | -0.017 [-0.12,0.085] |
|  | OTP | -10.645 | 6.106 | -1.743 | 0.081 | 0.265 | -0.091 [-0.194,0.011] |
|  | SCZ-AF | 1.249 | 8.562 | 0.146 | 0.884 | 0.942 | 0.008 [-0.095,0.110] |
|  | SCZ | -13.932 | 4.683 | -2.975 | **0.003** | **0.048** | -0.156 [-0.258,-0.053] |
| GC-ML-DG | BP I | -5.048 | 7.637 | -0.661 | 0.509 | 0.807 | -0.035 [-0.137,0.068] |
|  | BP II | 2.928 | 9.745 | 0.300 | 0.764 | 0.942 | 0.016 [-0.087,0.118] |
|  | OTP | -9.887 | 9.338 | -1.059 | 0.290 | 0.65 | -0.055 [-0.158,0.047] |
|  | SCZ-AF | -24.622 | 13.095 | -1.88 | 0.060 | 0.235 | -0.098 [-0.201,0.004] |
|  | SCZ | -22.862 | 7.162 | -3.192 | **0.001** | **0.047** | -0.167 [-0.27,-0.064] |
| HATA | BP I | 1.809 | 2.681 | 0.675 | 0.500 | 0.807 | 0.035 [-0.067,0.138] |
|  | BP II | 2.284 | 3.421 | 0.667 | 0.505 | 0.807 | 0.035 [-0.068,0.138] |
|  | OTP | -5.85 | 3.279 | -1.784 | 0.075 | 0.255 | -0.093 [-0.196,0.009] |
|  | SCZ-AF | -0.719 | 4.597 | -0.156 | 0.876 | 0.942 | -0.008 [-0.111,0.094] |
|  | SCZ | -4.546 | 2.514 | -1.808 | 0.071 | 0.255 | -0.095 [-0.197,0.008] |
| Tail | BP I | -5.981 | 21.907 | -0.273 | 0.785 | 0.942 | -0.014 [-0.117,0.088] |
|  | BP II | -40.374 | 27.952 | -1.444 | 0.149 | 0.393 | -0.076 [-0.178,0.027] |
|  | OTP | -52.955 | 26.786 | -1.977 | 0.048 | 0.235 | -0.103 [-0.206,-0.001] |
|  | SCZ-AF | -31.111 | 37.561 | -0.828 | 0.408 | 0.736 | -0.043 [-0.146,0.059] |
|  | SCZ | -34.9 | 20.542 | -1.699 | 0.090 | 0.277 | -0.089 [-0.192,0.014] |
| Fissure | BP I | -1.59 | 6.798 | -0.234 | 0.815 | 0.942 | -0.012 [-0.115,0.090] |
|  | BP II | -9.698 | 8.674 | -1.118 | 0.264 | 0.635 | -0.059 [-0.161,0.044] |
|  | OTP | 1.53 | 8.312 | 0.184 | 0.854 | 0.942 | 0.010 [-0.093,0.112] |
|  | SCZ-AF | -8.787 | 11.656 | -0.754 | 0.451 | 0.772 | -0.039 [-0.142,0.063] |
|  | SCZ | 2.958 | 6.374 | 0.464 | 0.643 | 0.889 | 0.024 [-0.078,0.127] |
| Molecular layer | BP I | -13.862 | 14.31 | -0.969 | 0.333 | 0.679 | -0.051 [-0.153,0.052] |
|  | BP II | 1.742 | 18.259 | 0.095 | 0.924 | 0.966 | 0.005 [-0.098,0.108] |
|  | OTP | -15.605 | 17.497 | -0.892 | 0.373 | 0.721 | -0.047 [-0.149,0.056] |
|  | SCZ-AF | -36.173 | 24.535 | -1.474 | 0.141 | 0.393 | -0.077 [-0.180,0.025] |
|  | SCZ | -40.724 | 13.418 | -3.035 | **0.002** | **0.048** | -0.159 [-0.262,-0.056] |
| Parasubiculum | BP I | -0.705 | 3.286 | -0.215 | 0.830 | 0.942 | -0.011 [-0.114,0.091] |
|  | BP II | 1.244 | 4.193 | 0.297 | 0.767 | 0.942 | 0.016 [-0.087,0.118] |
|  | OTP | -4.383 | 4.018 | -1.091 | 0.276 | 0.640 | -0.057 [-0.160,0.046] |
|  | SCZ-AF | -4.332 | 5.634 | -0.769 | 0.442 | 0.772 | -0.040 [-0.143,0.062] |
|  | SCZ | -6.448 | 3.081 | -2.093 | **0.037** | 0.235 | -0.110 [-0.212,-0.007] |
| Presubiculum | BP I | -10.504 | 10.142 | -1.036 | 0.301 | 0.651 | -0.054 [-0.157,0.048] |
|  | BP II | 0.79 | 12.941 | 0.061 | 0.951 | 0.966 | 0.003 [-0.099,0.106] |
|  | OTP | -24.811 | 12.401 | -2.001 | 0.046 | 0.235 | -0.105 [-0.207,-0.002] |
|  | SCZ-AF | -2.892 | 17.39 | -0.166 | 0.868 | 0.942 | -0.009 [-0.111,0.094] |
|  | SCZ | -20.837 | 9.51 | -2.191 | **0.029** | 0.232 | -0.115 [-0.217,-0.012] |
| Subiculum | BP I | -7.939 | 12.529 | -0.634 | 0.526 | 0.815 | -0.033 [-0.136,0.069] |
|  | BP II | -0.548 | 15.986 | -0.034 | 0.973 | 0.973 | -0.002 [-0.104,0.101] |
|  | OTP | -28.847 | 15.319 | -1.883 | 0.060 | 0.235 | -0.099 [-0.201,0.004] |
|  | SCZ-AF | -12.093 | 21.482 | -0.563 | 0.574 | 0.857 | -0.029 [-0.132,0.073] |
|  | SCZ | -27.488 | 11.748 | -2.34 | **0.019** | 0.180 | -0.122 [-0.225,-0.02] |
| Hippocampus | BP I | -69.745 | 82.587 | -0.844 | 0.399 | 0.736 | -0.044 [-0.147,0.058] |
|  | BP II | -30.153 | 105.379 | -0.286 | 0.775 | 0.942 | -0.015 [-0.118,0.088] |
|  | OTP | -166.404 | 100.982 | -1.648 | 0.100 | 0.294 | -0.086 [-0.189,0.016] |
|  | SCZ-AF | -203.323 | 141.603 | -1.436 | 0.151 | 0.393 | -0.075 [-0.178,0.027] |
|  | SCZ | -251.581 | 77.442 | -3.249 | **0.001** | **0.047** | -0.170 [-0.273,-0.067] |

Statistical results are based on multiple linear regression (adjusted for age, age^2^, intracranial volume). Abbreviation: BP = bipolar, SCZ = schizophrenia, SCZ-AF = schizoaffective disorder, OPD = other psychotic disorders, CA = cornu ammonis, GC-ML-DG = granule cells in the molecular layer of the dentate gyrus, HATA = hippocampal-amygdaloid transition area, FDR = false discover rate, CI = confidence Interval. Significant results before and after FDR-correction for multiple comparison are highlighted in bold.

**Table S7| Associations between hippocampal volumes and medication in bipolar disorders (BD) and schizophrenia spectrum disorders (SZ).**

| **Subfields** | **Diagnosis** | **Test** | **Variable** | **tvalue** | **pvalue** | **pFDR** |
| --- | --- | --- | --- | --- | --- | --- |
| CA1 | BD | Lithium, status | main | 1.157 | 0.248 | 0.979 |
|  | BD | Lithium, status | interaction | -0.418 | 0.676 | 0.979 |
|  | BD | Lithium, serum | main | 1.577 | 0.116 | 0.975 |
|  | BD | Lithium, serum | interaction | -0.217 | 0.828 | 0.985 |
|  | BD | Antipsychotics, DDD | main | 0.493 | 0.623 | 0.979 |
|  | BD | AP, DDD | interaction | 1.415 | 0.159 | 0.975 |
|  | BD | AD, DDD | main | -0.432 | 0.666 | 0.979 |
|  | BD | AD, DDD | interaction | 1.401 | 0.164 | 0.975 |
|  | BD | AE, DDD | main | 0.158 | 0.875 | 0.994 |
|  | BD | AE, DDD | interaction | 0.048 | 0.962 | 0.994 |
|  | SZ | AP, DDD | main | 1.037 | 0.300 | 0.979 |
|  | SZ | AP, DDD | interaction | 0.825 | 0.410 | 0.979 |
|  | SZ | AD, DDD | main | -0.409 | 0.683 | 0.979 |
|  | SZ | AD, DDD | interaction | 1.559 | 0.121 | 0.975 |
|  | SZ | AE, DDD | main | -0.556 | 0.582 | 0.979 |
|  | SZ | AE, DDD | interaction | 0.819 | 0.418 | 0.979 |
| CA3 | BD | Lithium, status | main | 1.806 | 0.072 | 0.975 |
|  | BD | Lithium, status | interaction | 0.077 | 0.939 | 0.994 |
|  | BD | Lithium, serum | main | 1.789 | 0.075 | 0.975 |
|  | BD | Lithium, serum | interaction | -0.008 | 0.994 | 0.994 |
|  | BD | AP, DDD | main | -0.766 | 0.445 | 0.979 |
|  | BD | AP, DDD | interaction | 0.499 | 0.619 | 0.979 |
|  | BD | AD, DDD | main | -0.424 | 0.673 | 0.979 |
|  | BD | AD, DDD | interaction | 0.977 | 0.331 | 0.979 |
|  | BD | AE, DDD | main | -0.322 | 0.748 | 0.979 |
|  | BD | AE, DDD | interaction | -0.506 | 0.614 | 0.979 |
|  | SZ | AP, DDD | main | 0.556 | 0.578 | 0.979 |
|  | SZ | AP, DDD | interaction | -0.616 | 0.538 | 0.979 |
|  | SZ | AD, DDD | main | 0.821 | 0.413 | 0.979 |
|  | SZ | AD, DDD | interaction | 0.994 | 0.322 | 0.979 |
|  | SZ | AE, DDD | main | 1.523 | 0.136 | 0.975 |
|  | SZ | AE, DDD | interaction | 0.029 | 0.977 | 0.994 |
| CA4 | BD | Lithium, status | main | 1.321 | 0.187 | 0.979 |
|  | BD | Lithium, status | interaction | -0.703 | 0.483 | 0.979 |
|  | BD | Lithium, serum | main | 1.658 | 0.098 | 0.975 |
|  | BD | Lithium, serum | interaction | -0.429 | 0.668 | 0.979 |
|  | BD | AP, DDD | main | -0.085 | 0.932 | 0.994 |
|  | BD | AP, DDD | interaction | 1.130 | 0.260 | 0.979 |
|  | BD | AD, DDD | main | 0.022 | 0.982 | 0.994 |
|  | BD | AD, DDD | interaction | 1.171 | 0.244 | 0.979 |
|  | BD | AE, DDD | main | -0.226 | 0.821 | 0.985 |
|  | BD | AE, DDD | interaction | -0.419 | 0.676 | 0.979 |
|  | SZ | AP, DDD | main | 0.907 | 0.365 | 0.979 |
|  | SZ | AP, DDD | interaction | 0.068 | 0.946 | 0.994 |
|  | SZ | AD, DDD | main | 0.497 | 0.620 | 0.979 |
|  | SZ | AD, DDD | interaction | 0.855 | 0.394 | 0.979 |
|  | SZ | AE, DDD | main | 0.435 | 0.666 | 0.979 |
|  | SZ | AE, DDD | interaction | 0.584 | 0.563 | 0.979 |
| Fimbria | BD | Lithium, status | main | 0.253 | 0.800 | 0.984 |
|  | BD | Lithium, status | interaction | 0.327 | 0.744 | 0.979 |
|  | BD | Lithium, serum | main | 0.655 | 0.513 | 0.979 |
|  | BD | Lithium, serum | interaction | 0.647 | 0.518 | 0.979 |
|  | BD | AP, DDD | main | 2.100 | **0.037** | 0.975 |
|  | BD | AP, DDD | interaction | -0.802 | 0.424 | 0.979 |
|  | BD | AD, DDD | main | -0.695 | 0.489 | 0.979 |
|  | BD | AD, DDD | interaction | 1.100 | 0.274 | 0.979 |
|  | BD | AE, DDD | main | -1.247 | 0.215 | 0.979 |
|  | BD | AE, DDD | interaction | -0.830 | 0.408 | 0.979 |
|  | SZ | AP, DDD | main | -0.079 | 0.937 | 0.994 |
|  | SZ | AP, DDD | interaction | -0.451 | 0.652 | 0.979 |
|  | SZ | AD, DDD | main | -2.025 | **0.045** | 0.975 |
|  | SZ | AD, DDD | interaction | -0.023 | 0.982 | 0.994 |
|  | SZ | AE, DDD | main | -1.141 | 0.261 | 0.979 |
|  | SZ | AE, DDD | interaction | 0.319 | 0.751 | 0.979 |
| GC-ML-DG | BD | Lithium, status | main | 1.399 | 0.163 | 0.975 |
|  | BD | Lithium, status | interaction | -0.643 | 0.521 | 0.979 |
|  | BD | Lithium, serum | main | 1.701 | 0.090 | 0.975 |
|  | BD | Lithium, serum | interaction | -0.301 | 0.764 | 0.979 |
|  | BD | AP, DDD | main | 0.301 | 0.764 | 0.979 |
|  | BD | AP, DDD | interaction | 1.096 | 0.275 | 0.979 |
|  | BD | AD, DDD | main | 0.033 | 0.974 | 0.994 |
|  | BD | AD, DDD | interaction | 1.264 | 0.209 | 0.979 |
|  | BD | AE, DDD | main | -0.284 | 0.777 | 0.979 |
|  | BD | AE, DDD | interaction | -0.360 | 0.720 | 0.979 |
|  | SZ | AP, DDD | main | 0.967 | 0.334 | 0.979 |
|  | SZ | AP, DDD | interaction | 0.032 | 0.975 | 0.994 |
|  | SZ | AD, DDD | main | 0.256 | 0.799 | 0.984 |
|  | SZ | AD, DDD | interaction | 0.913 | 0.363 | 0.979 |
|  | SZ | AE, DDD | main | 0.168 | 0.868 | 0.994 |
|  | SZ | AE, DDD | interaction | 0.537 | 0.594 | 0.979 |
| HATA | BD | Lithium, status | main | 2.051 | **0.041** | 0.975 |
|  | BD | Lithium, status | interaction | 1.783 | 0.076 | 0.975 |
|  | BD | Lithium, serum | main | 2.681 | **0.008** | 0.975 |
|  | BD | Lithium, serum | interaction | 1.845 | 0.066 | 0.975 |
|  | BD | AP, DDD | main | 1.650 | 0.101 | 0.975 |
|  | BD | AP, DDD | interaction | 0.475 | 0.636 | 0.979 |
|  | BD | AD, DDD | main | 1.425 | 0.157 | 0.975 |
|  | BD | AD, DDD | interaction | 0.629 | 0.531 | 0.979 |
|  | BD | AE, DDD | main | -0.916 | 0.362 | 0.979 |
|  | BD | AE, DDD | interaction | -0.205 | 0.838 | 0.987 |
|  | SZ | AP, DDD | main | 0.222 | 0.824 | 0.985 |
|  | SZ | AP, DDD | interaction | -1.211 | 0.227 | 0.979 |
|  | SZ | AD, DDD | main | -1.213 | 0.227 | 0.979 |
|  | SZ | AD, DDD | interaction | 0.387 | 0.699 | 0.979 |
|  | SZ | AE, DDD | main | -0.560 | 0.579 | 0.979 |
|  | SZ | AE, DDD | interaction | 0.560 | 0.579 | 0.979 |
| Fissure | BD | Lithium, status | main | -0.255 | 0.799 | 0.984 |
|  | BD | Lithium, status | interaction | -0.497 | 0.619 | 0.979 |
|  | BD | Lithium, serum | main | -0.727 | 0.468 | 0.979 |
|  | BD | Lithium, serum | interaction | -0.010 | 0.992 | 0.994 |
|  | BD | AP, DDD | main | -2.145 | **0.034** | 0.975 |
|  | BD | AP, DDD | interaction | 1.504 | 0.135 | 0.975 |
|  | BD | AD, DDD | main | -0.286 | 0.776 | 0.979 |
|  | BD | AD, DDD | interaction | 0.981 | 0.329 | 0.979 |
|  | BD | AE, DDD | main | 2.230 | **0.028** | 0.975 |
|  | BD | AE, DDD | interaction | -1.694 | 0.093 | 0.975 |
|  | SZ | AP, DDD | main | -1.135 | 0.257 | 0.979 |
|  | SZ | AP, DDD | interaction | 1.268 | 0.205 | 0.979 |
|  | SZ | AD, DDD | main | 0.908 | 0.366 | 0.979 |
|  | SZ | AD, DDD | interaction | -0.131 | 0.896 | 0.994 |
|  | SZ | AE, DDD | main | -2.284 | **0.028** | 0.975 |
|  | SZ | AE, DDD | interaction | 0.505 | 0.617 | 0.979 |
| Tail | BD | Lithium, status | main | 0.500 | 0.618 | 0.979 |
|  | BD | Lithium, status | interaction | -0.859 | 0.391 | 0.979 |
|  | BD | Lithium, serum | main | 1.434 | 0.153 | 0.975 |
|  | BD | Lithium, serum | interaction | -0.714 | 0.476 | 0.979 |
|  | BD | AP, DDD | main | -0.493 | 0.623 | 0.979 |
|  | BD | AP, DDD | interaction | 1.074 | 0.284 | 0.979 |
|  | BD | AD, DDD | main | -0.017 | 0.987 | 0.994 |
|  | BD | AD, DDD | interaction | 0.402 | 0.689 | 0.979 |
|  | BD | AE, DDD | main | -0.565 | 0.573 | 0.979 |
|  | BD | AE, DDD | interaction | 0.026 | 0.979 | 0.994 |
|  | SZ | AP, DDD | main | 0.059 | 0.953 | 0.994 |
|  | SZ | AP, DDD | interaction | -0.376 | 0.707 | 0.979 |
|  | SZ | AD, DDD | main | 0.202 | 0.840 | 0.987 |
|  | SZ | AD, DDD | interaction | 0.516 | 0.607 | 0.979 |
|  | SZ | AE, DDD | main | -1.641 | 0.109 | 0.975 |
|  | SZ | AE, DDD | interaction | -0.695 | 0.491 | 0.979 |
| Molecular layer | BD | Lithium, status | main | 1.127 | 0.261 | 0.979 |
|  | BD | Lithium, status | interaction | -0.984 | 0.326 | 0.979 |
|  | BD | Lithium, serum | main | 1.952 | 0.052 | 0.975 |
|  | BD | Lithium, serum | interaction | -0.487 | 0.627 | 0.979 |
|  | BD | AP, DDD | main | -0.011 | 0.991 | 0.994 |
|  | BD | AP, DDD | interaction | 1.163 | 0.247 | 0.979 |
|  | BD | AD, DDD | main | -0.373 | 0.710 | 0.979 |
|  | BD | AD, DDD | interaction | 1.634 | 0.105 | 0.975 |
|  | BD | AE, DDD | main | -0.148 | 0.883 | 0.994 |
|  | BD | AE, DDD | interaction | 0.132 | 0.895 | 0.994 |
|  | SZ | AP, DDD | main | 1.127 | 0.260 | 0.979 |
|  | SZ | AP, DDD | interaction | 0.556 | 0.579 | 0.979 |
|  | SZ | AD, DDD | main | -0.531 | 0.596 | 0.979 |
|  | SZ | AD, DDD | interaction | 1.610 | 0.110 | 0.975 |
|  | SZ | AE, DDD | main | -0.587 | 0.560 | 0.979 |
|  | SZ | AE, DDD | interaction | 0.521 | 0.605 | 0.979 |
| Parasubiculum | BD | Lithium, status | main | -0.898 | 0.370 | 0.979 |
|  | BD | Lithium, status | interaction | -0.102 | 0.919 | 0.994 |
|  | BD | Lithium, serum | main | 0.219 | 0.827 | 0.985 |
|  | BD | Lithium, serum | interaction | 0.183 | 0.855 | 0.989 |
|  | BD | AP, DDD | main | -0.865 | 0.388 | 0.979 |
|  | BD | AP, DDD | interaction | 0.789 | 0.431 | 0.979 |
|  | BD | AD, DDD | main | 0.862 | 0.391 | 0.979 |
|  | BD | AD, DDD | interaction | 0.390 | 0.698 | 0.979 |
|  | BD | AE, DDD | main | -0.222 | 0.825 | 0.985 |
|  | BD | AE, DDD | interaction | 0.652 | 0.516 | 0.979 |
|  | SZ | AP, DDD | main | -0.019 | 0.985 | 0.994 |
|  | SZ | AP, DDD | interaction | 1.451 | 0.148 | 0.975 |
|  | SZ | AD, DDD | main | -0.463 | 0.644 | 0.979 |
|  | SZ | AD, DDD | interaction | -0.252 | 0.802 | 0.984 |
|  | SZ | AE, DDD | main | 1.125 | 0.267 | 0.979 |
|  | SZ | AE, DDD | interaction | 0.091 | 0.928 | 0.994 |
| Presubiculum | BD | Lithium, status | main | -0.554 | 0.580 | 0.979 |
|  | BD | Lithium, status | interaction | -0.766 | 0.444 | 0.979 |
|  | BD | Lithium, serum | main | 1.241 | 0.216 | 0.979 |
|  | BD | Lithium, serum | interaction | 0.470 | 0.639 | 0.979 |
|  | BD | AP, DDD | main | -0.729 | 0.467 | 0.979 |
|  | BD | AP, DDD | interaction | 1.507 | 0.134 | 0.975 |
|  | BD | AD, DDD | main | 0.687 | 0.494 | 0.979 |
|  | BD | AD, DDD | interaction | 1.282 | 0.203 | 0.979 |
|  | BD | AE, DDD | main | -0.631 | 0.529 | 0.979 |
|  | BD | AE, DDD | interaction | -0.973 | 0.333 | 0.979 |
|  | SZ | AP, DDD | main | -0.303 | 0.762 | 0.979 |
|  | SZ | AP, DDD | interaction | 0.924 | 0.356 | 0.979 |
|  | SZ | AD, DDD | main | -1.239 | 0.218 | 0.979 |
|  | SZ | AD, DDD | interaction | 0.974 | 0.332 | 0.979 |
|  | SZ | AE, DDD | main | -1.517 | 0.137 | 0.975 |
|  | SZ | AE, DDD | interaction | 0.342 | 0.734 | 0.979 |
| Subiculum | BD | Lithium, status | main | -0.144 | 0.886 | 0.994 |
|  | BD | Lithium, status | interaction | -0.994 | 0.321 | 0.979 |
|  | BD | Lithium, serum | main | 1.040 | 0.299 | 0.979 |
|  | BD | LIT, serum | interaction | -0.385 | 0.701 | 0.979 |
|  | BD | AP, DDD | main | -0.249 | 0.804 | 0.984 |
|  | BD | AP, DDD | interaction | 0.065 | 0.948 | 0.994 |
|  | BD | AD, DDD | main | -0.715 | 0.476 | 0.979 |
|  | BD | AD, DDD | interaction | 2.058 | **0.042** | 0.975 |
|  | BD | AE, DDD | main | 0.182 | 0.856 | 0.989 |
|  | BD | AE, DDD | interaction | -0.505 | 0.615 | 0.979 |
|  | SZ | AP, DDD | main | 0.051 | 0.960 | 0.994 |
|  | SZ | AP, DDD | interaction | 0.710 | 0.478 | 0.979 |
|  | SZ | AD, DDD | main | -0.828 | 0.409 | 0.979 |
|  | SZ | AD, DDD | interaction | 1.298 | 0.197 | 0.979 |
|  | SZ | AE, DDD | main | -1.327 | 0.192 | 0.979 |
|  | SZ | AE, DDD | interaction | 0.320 | 0.751 | 0.979 |
| Hippocampus | BD | Lithium, status | main | 0.989 | 0.323 | 0.979 |
|  | BD | Lithium, status | interaction | -0.784 | 0.434 | 0.979 |
|  | BD | Lithium, serum | main | 2.043 | 0.042 | 0.975 |
|  | BD | Lithium, serum | interaction | -0.284 | 0.777 | 0.979 |
|  | BD | AP, DDD | main | -0.057 | 0.955 | 0.994 |
|  | BD | AP, DDD | interaction | 1.253 | 0.212 | 0.979 |
|  | BD | AD, DDD | main | -0.183 | 0.855 | 0.989 |
|  | BD | AD, DDD | interaction | 1.616 | 0.109 | 0.975 |
|  | BD | AE, DDD | main | -0.395 | 0.694 | 0.979 |
|  | BD | AE, DDD | interaction | -0.321 | 0.749 | 0.979 |
|  | SZ | AP, DDD | main | 0.660 | 0.510 | 0.979 |
|  | SZ | AP, DDD | interaction | 0.347 | 0.728 | 0.979 |
|  | SZ | AD, DDD | main | -0.450 | 0.654 | 0.979 |
|  | SZ | AD, DDD | interaction | 1.411 | 0.161 | 0.975 |
|  | SZ | AE, DDD | main | -0.878 | 0.385 | 0.979 |
|  | SZ | AE, DDD | interaction | 0.330 | 0.743 | 0.979 |

Main effects are based on regression models, adjusted for sex, age, age^2^ and intracranial volume. Interaction effects are based on regression models with a sex-by-clinical measures interaction term, adjusted for age, age^2^ and intracranial volume. Abbreviation: CA = cornu ammonis, GC-ML-DG = granule cells in the molecular layer of the dentate gyrus, HATA = hippocampal-amygdaloid transition area, DDD = daily defined dose, FDR = false discover rate. Significant results are highlighted in bold.

**Table S8| Associations between hippocampal volumes and clinical measures in bipolar disorders (BD) and schizophrenia spectrum disorders (SZ).**

| **Subfields** | **Diagnosis** | **Test** | **Effect** | **tvalue** | **pvalue** | **pFDR** |
| --- | --- | --- | --- | --- | --- | --- |
| CA1 | BD | GAF, F | main | 0.622 | 0.534 | 0.979 |
|  | BD | GAF, F | interaction | -0.974 | 0.331 | 0.955 |
|  | BD | GAF, S | main | -0.009 | 0.993 | 0.998 |
|  | BD | GAF, S | interaction | 1.223 | 0.222 | 0.878 |
|  | BD | IDS | main | -0.126 | 0.900 | 0.979 |
|  | BD | IDS | interaction | -0.109 | 0.913 | 0.979 |
|  | BD | YMRS | main | -0.749 | 0.454 | 0.979 |
|  | BD | YMRS | interaction | -1.596 | 0.112 | 0.767 |
|  | BD | AOO | main | -2.738 | **0.007** | 0.227 |
|  | BD | AOO | interaction | -0.476 | 0.635 | 0.979 |
|  | BD | DOI | main | -1.068 | 0.286 | 0.955 |
|  | BD | DOI | interaction | -2.183 | **0.030** | 0.607 |
|  | SZ | GAF, F | main | 0.038 | 0.970 | 0.989 |
|  | SZ | GAF, F | interaction | 0.816 | 0.415 | 0.979 |
|  | SZ | GAF, S | main | 0.488 | 0.626 | 0.979 |
|  | SZ | GAF, S | interaction | 0.407 | 0.685 | 0.979 |
|  | SZ | score, n | main | -0.247 | 0.805 | 0.979 |
|  | SZ | score, n | interaction | -0.371 | 0.711 | 0.979 |
|  | SZ | score, p | main | -0.124 | 0.901 | 0.979 |
|  | SZ | score, p | interaction | -0.320 | 0.749 | 0.979 |
|  | SZ | AOO | main | 0.727 | 0.467 | 0.979 |
|  | SZ | AOO | interaction | -0.439 | 0.661 | 0.979 |
|  | SZ | DOI | main | -0.211 | 0.833 | 0.979 |
|  | SZ | DOI | interaction | -0.253 | 0.801 | 0.979 |
| CA3 | BD | GAF, F | main | 1.279 | 0.202 | 0.843 |
|  | BD | GAF, F | interaction | 0.540 | 0.589 | 0.979 |
|  | BD | GAF, S | main | 0.534 | 0.594 | 0.979 |
|  | BD | GAF, S | interaction | 1.655 | 0.099 | 0.702 |
|  | BD | IDS | main | -0.029 | 0.977 | 0.989 |
|  | BD | IDS | interaction | -0.810 | 0.418 | 0.979 |
|  | BD | YMRS | main | -0.318 | 0.751 | 0.979 |
|  | BD | YMRS | interaction | -0.578 | 0.563 | 0.979 |
|  | BD | AOO | main | -2.143 | **0.033** | 0.607 |
|  | BD | AOO | interaction | -0.433 | 0.666 | 0.979 |
|  | BD | DOI | main | -0.444 | 0.657 | 0.979 |
|  | BD | DOI | interaction | 0.217 | 0.828 | 0.979 |
|  | SZ | GAF, F | main | 0.803 | 0.422 | 0.979 |
|  | SZ | GAF, F | interaction | 0.793 | 0.428 | 0.979 |
|  | SZ | GAF, S | main | 1.919 | 0.056 | 0.607 |
|  | SZ | GAF, S | interaction | 0.122 | 0.903 | 0.979 |
|  | SZ | score, n | main | 0.114 | 0.909 | 0.979 |
|  | SZ | score, n | interaction | -0.875 | 0.382 | 0.979 |
|  | SZ | score, p | main | -1.957 | 0.051 | 0.607 |
|  | SZ | score, p | interaction | -1.254 | 0.211 | 0.854 |
|  | SZ | AOO | main | 1.892 | 0.059 | 0.607 |
|  | SZ | AOO | interaction | -0.206 | 0.837 | 0.979 |
|  | SZ | DOI | main | 0.420 | 0.675 | 0.979 |
|  | SZ | DOI | interaction | -0.467 | 0.641 | 0.979 |
| CA4 | BD | GAF, F | main | 1.407 | 0.160 | 0.828 |
|  | BD | GAF, F | interaction | -0.132 | 0.895 | 0.979 |
|  | BD | GAF, S | main | 0.364 | 0.716 | 0.979 |
|  | BD | GAF, S | interaction | 1.458 | 0.146 | 0.827 |
|  | BD | IDS | main | 0.405 | 0.686 | 0.979 |
|  | BD | IDS | interaction | -0.148 | 0.882 | 0.979 |
|  | BD | YMRS | main | -0.162 | 0.872 | 0.979 |
|  | BD | YMRS | interaction | -0.822 | 0.412 | 0.979 |
|  | BD | AOO | main | -2.467 | **0.014** | 0.402 |
|  | BD | AOO | interaction | -1.231 | 0.219 | 0.877 |
|  | BD | DOI | main | 0.007 | 0.994 | 0.998 |
|  | BD | DOI | interaction | 0.278 | 0.781 | 0.979 |
|  | SZ | GAF, F | main | 0.498 | 0.619 | 0.979 |
|  | SZ | GAF, F | interaction | 0.980 | 0.328 | 0.955 |
|  | SZ | GAF, S | main | 1.333 | 0.183 | 0.828 |
|  | SZ | GAF, S | interaction | 0.407 | 0.685 | 0.979 |
|  | SZ | score, n | main | -0.357 | 0.722 | 0.979 |
|  | SZ | score, n | interaction | -1.283 | 0.200 | 0.843 |
|  | SZ | score, p | main | -1.081 | 0.280 | 0.955 |
|  | SZ | score, p | interaction | -1.523 | 0.128 | 0.774 |
|  | SZ | AOO | main | 1.341 | 0.181 | 0.828 |
|  | SZ | AOO | interaction | -0.336 | 0.737 | 0.979 |
|  | SZ | DOI | main | 0.465 | 0.642 | 0.979 |
|  | SZ | DOI | interaction | 0.225 | 0.822 | 0.979 |
| Fimbria | BD | GAF, F | main | -1.779 | 0.076 | 0.617 |
|  | BD | GAF, F | interaction | -0.808 | 0.420 | 0.979 |
|  | BD | GAF, S | main | -0.599 | 0.549 | 0.979 |
|  | BD | GAF, S | interaction | -0.507 | 0.613 | 0.979 |
|  | BD | IDS | main | -0.294 | 0.769 | 0.979 |
|  | BD | IDS | interaction | 1.290 | 0.198 | 0.843 |
|  | BD | YMRS | main | 1.350 | 0.178 | 0.828 |
|  | BD | YMRS | interaction | -0.230 | 0.819 | 0.979 |
|  | BD | AOO | main | -3.195 | **0.002** | 0.098 |
|  | BD | AOO | interaction | -1.789 | 0.075 | 0.617 |
|  | BD | DOI | main | -1.823 | 0.069 | 0.617 |
|  | BD | DOI | interaction | -1.904 | 0.058 | 0.607 |
|  | SZ | GAF, F | main | -0.755 | 0.451 | 0.979 |
|  | SZ | GAF, F | interaction | -0.956 | 0.340 | 0.955 |
|  | SZ | GAF, S | main | -0.114 | 0.910 | 0.979 |
|  | SZ | GAF, S | interaction | -1.268 | 0.205 | 0.843 |
|  | SZ | score, n | main | -0.539 | 0.590 | 0.979 |
|  | SZ | score, n | interaction | 1.921 | 0.055 | 0.607 |
|  | SZ | score, p | main | 0.539 | 0.590 | 0.979 |
|  | SZ | score, p | interaction | 0.631 | 0.528 | 0.979 |
|  | SZ | AOO | main | -1.139 | 0.255 | 0.949 |
|  | SZ | AOO | interaction | 0.234 | 0.815 | 0.979 |
|  | SZ | DOI | main | -1.477 | 0.140 | 0.812 |
|  | SZ | DOI | interaction | 0.502 | 0.616 | 0.979 |
| GC-ML-DG | BD | GAF, F | main | 1.380 | 0.169 | 0.828 |
|  | BD | GAF, F | interaction | 0.055 | 0.956 | 0.989 |
|  | BD | GAF, S | main | 0.286 | 0.775 | 0.979 |
|  | BD | GAF, S | interaction | 1.555 | 0.121 | 0.767 |
|  | BD | IDS | main | 0.119 | 0.905 | 0.979 |
|  | BD | IDS | interaction | -0.170 | 0.865 | 0.979 |
|  | BD | YMRS | main | -0.200 | 0.842 | 0.979 |
|  | BD | YMRS | interaction | -1.165 | 0.245 | 0.944 |
|  | BD | AOO | main | -3.169 | **0.002** | 0.098 |
|  | BD | AOO | interaction | -1.547 | 0.123 | 0.767 |
|  | BD | DOI | main | -0.123 | 0.902 | 0.979 |
|  | BD | DOI | interaction | -0.057 | 0.954 | 0.989 |
|  | SZ | GAF, F | main | 0.411 | 0.681 | 0.979 |
|  | SZ | GAF, F | interaction | 0.995 | 0.320 | 0.955 |
|  | SZ | GAF, S | main | 1.127 | 0.261 | 0.955 |
|  | SZ | GAF, S | interaction | 0.523 | 0.601 | 0.979 |
|  | SZ | score, n | main | -0.330 | 0.742 | 0.979 |
|  | SZ | score, n | interaction | -1.360 | 0.175 | 0.828 |
|  | SZ | score, p | main | -0.785 | 0.433 | 0.979 |
|  | SZ | score, p | interaction | -1.276 | 0.203 | 0.843 |
|  | SZ | AOO | main | 1.034 | 0.302 | 0.955 |
|  | SZ | AOO | interaction | -0.535 | 0.593 | 0.979 |
|  | SZ | DOI | main | 0.033 | 0.974 | 0.989 |
|  | SZ | DOI | interaction | 0.500 | 0.617 | 0.979 |
| HATA | BD | GAF, F | main | 0.954 | 0.341 | 0.955 |
|  | BD | GAF, F | interaction | 2.417 | **0.016** | 0.423 |
|  | BD | GAF, S | main | 0.083 | 0.934 | 0.989 |
|  | BD | GAF, S | interaction | 2.017 | **0.045** | 0.607 |
|  | BD | IDS | main | 0.310 | 0.756 | 0.979 |
|  | BD | IDS | interaction | -0.822 | 0.412 | 0.979 |
|  | BD | YMRS | main | -1.371 | 0.171 | 0.828 |
|  | BD | YMRS | interaction | -1.336 | 0.183 | 0.828 |
|  | BD | AOO | main | -3.135 | **0.002** | 0.098 |
|  | BD | AOO | interaction | -1.021 | 0.308 | 0.955 |
|  | BD | DOI | main | 0.504 | 0.614 | 0.979 |
|  | BD | DOI | interaction | -0.806 | 0.421 | 0.979 |
|  | SZ | GAF, F | main | -0.054 | 0.957 | 0.989 |
|  | SZ | GAF, F | interaction | 0.318 | 0.751 | 0.979 |
|  | SZ | GAF, S | main | 0.101 | 0.920 | 0.979 |
|  | SZ | GAF, S | interaction | -0.333 | 0.739 | 0.979 |
|  | SZ | score, n | main | -0.994 | 0.321 | 0.955 |
|  | SZ | score, n | interaction | -0.246 | 0.806 | 0.979 |
|  | SZ | score, p | main | -0.133 | 0.895 | 0.979 |
|  | SZ | score, p | interaction | 0.372 | 0.710 | 0.979 |
|  | SZ | AOO | main | 0.343 | 0.732 | 0.979 |
|  | SZ | AOO | interaction | -0.716 | 0.474 | 0.979 |
|  | SZ | DOI | main | -0.103 | 0.918 | 0.979 |
|  | SZ | DOI | interaction | 0.599 | 0.549 | 0.979 |
| Fissure | BD | GAF, F | main | -0.075 | 0.940 | 0.989 |
|  | BD | GAF, F | interaction | -0.590 | 0.556 | 0.979 |
|  | BD | GAF, S | main | 0.060 | 0.952 | 0.989 |
|  | BD | GAF, S | interaction | 1.728 | 0.085 | 0.617 |
|  | BD | IDS | main | -0.216 | 0.829 | 0.979 |
|  | BD | IDS | interaction | -0.175 | 0.861 | 0.979 |
|  | BD | YMRS | main | 0.547 | 0.585 | 0.979 |
|  | BD | YMRS | interaction | -1.872 | 0.062 | 0.607 |
|  | BD | AOO | main | 4.149 | **4.32e-05** | **0.007** |
|  | BD | AOO | interaction | 2.272 | **0.024** | 0.531 |
|  | BD | DOI | main | 3.713 | **2.43e-04** | **0.025** |
|  | BD | DOI | interaction | -2.325 | **0.021** | 0.498 |
|  | SZ | GAF, F | main | -1.004 | 0.316 | 0.955 |
|  | SZ | GAF, F | interaction | 1.012 | 0.312 | 0.955 |
|  | SZ | GAF, S | main | -1.445 | 0.149 | 0.828 |
|  | SZ | GAF, S | interaction | 0.314 | 0.754 | 0.979 |
|  | SZ | score, n | main | 0.716 | 0.475 | 0.979 |
|  | SZ | score, n | interaction | -0.063 | 0.949 | 0.989 |
|  | SZ | score, p | main | 0.951 | 0.342 | 0.955 |
|  | SZ | score, p | interaction | -1.364 | 0.173 | 0.828 |
|  | SZ | AOO | main | 3.030 | **0.003** | 0.115 |
|  | SZ | AOO | interaction | 0.799 | 0.425 | 0.979 |
|  | SZ | DOI | main | 4.432 | **1.18e-05** | **0.004** |
|  | SZ | DOI | interaction | 0.419 | 0.676 | 0.979 |
| Tail | BD | GAF, F | main | 0.883 | 0.378 | 0.979 |
|  | BD | GAF, F | interaction | -0.357 | 0.722 | 0.979 |
|  | BD | GAF, S | main | 1.287 | 0.199 | 0.843 |
|  | BD | GAF, S | interaction | 0.809 | 0.419 | 0.979 |
|  | BD | IDS | main | 0.704 | 0.482 | 0.979 |
|  | BD | IDS | interaction | -0.300 | 0.764 | 0.979 |
|  | BD | YMRS | main | 0.837 | 0.403 | 0.979 |
|  | BD | YMRS | interaction | -1.100 | 0.272 | 0.955 |
|  | BD | AOO | main | -0.548 | 0.584 | 0.979 |
|  | BD | AOO | interaction | -1.003 | 0.317 | 0.955 |
|  | BD | DOI | main | -1.356 | 0.176 | 0.828 |
|  | BD | DOI | interaction | -1.879 | 0.061 | 0.607 |
|  | SZ | GAF, F | main | 2.031 | **0.043** | 0.607 |
|  | SZ | GAF, F | interaction | 0.513 | 0.608 | 0.979 |
|  | SZ | GAF, S | main | 1.882 | 0.060 | 0.607 |
|  | SZ | GAF, S | interaction | -0.286 | 0.775 | 0.979 |
|  | SZ | score, n | main | -1.383 | 0.167 | 0.828 |
|  | SZ | score, n | interaction | -0.370 | 0.712 | 0.979 |
|  | SZ | score, p | main | -1.558 | 0.120 | 0.767 |
|  | SZ | score, p | interaction | 0.221 | 0.825 | 0.979 |
|  | SZ | AOO | main | 0.320 | 0.749 | 0.979 |
|  | SZ | AOO | interaction | -0.907 | 0.365 | 0.979 |
|  | SZ | DOI | main | 0.752 | 0.452 | 0.979 |
|  | SZ | DOI | interaction | -0.021 | 0.984 | 0.993 |
| Molecular layer | BD | GAF, F | main | 1.004 | 0.316 | 0.955 |
|  | BD | GAF, F | interaction | -0.960 | 0.338 | 0.955 |
|  | BD | GAF, S | main | 0.434 | 0.665 | 0.979 |
|  | BD | GAF, S | interaction | 1.112 | 0.267 | 0.955 |
|  | BD | IDS | main | 0.223 | 0.823 | 0.979 |
|  | BD | IDS | interaction | 0.238 | 0.812 | 0.979 |
|  | BD | YMRS | main | -0.549 | 0.583 | 0.979 |
|  | BD | YMRS | interaction | -1.512 | 0.132 | 0.774 |
|  | BD | AOO | main | -2.844 | **0.005** | 0.186 |
|  | BD | AOO | interaction | -0.767 | 0.444 | 0.979 |
|  | BD | DOI | main | -0.264 | 0.792 | 0.979 |
|  | BD | DOI | interaction | -1.765 | 0.079 | 0.617 |
|  | SZ | GAF, F | main | 0.044 | 0.965 | 0.989 |
|  | SZ | GAF, F | interaction | 0.841 | 0.401 | 0.979 |
|  | SZ | GAF, S | main | 0.576 | 0.565 | 0.979 |
|  | SZ | GAF, S | interaction | 0.563 | 0.574 | 0.979 |
|  | SZ | score, n | main | -0.107 | 0.915 | 0.979 |
|  | SZ | score, n | interaction | -0.768 | 0.443 | 0.979 |
|  | SZ | score, p | main | -0.161 | 0.872 | 0.979 |
|  | SZ | score, p | interaction | -0.601 | 0.548 | 0.979 |
|  | SZ | AOO | main | 0.896 | 0.371 | 0.979 |
|  | SZ | AOO | interaction | -0.632 | 0.528 | 0.979 |
|  | SZ | DOI | main | 0.002 | 0.999 | 0.999 |
|  | SZ | DOI | interaction | 0.438 | 0.661 | 0.979 |
| Parasubiculum | BD | GAF, F | main | 0.539 | 0.590 | 0.979 |
|  | BD | GAF, F | interaction | -0.538 | 0.591 | 0.979 |
|  | BD | GAF, S | main | -0.127 | 0.899 | 0.979 |
|  | BD | GAF, S | interaction | -0.482 | 0.630 | 0.979 |
|  | BD | IDS | main | 1.742 | 0.083 | 0.617 |
|  | BD | IDS | interaction | 0.411 | 0.681 | 0.979 |
|  | BD | YMRS | main | -0.392 | 0.695 | 0.979 |
|  | BD | YMRS | interaction | -0.663 | 0.508 | 0.979 |
|  | BD | AOO | main | -1.752 | 0.081 | 0.617 |
|  | BD | AOO | interaction | -0.356 | 0.722 | 0.979 |
|  | BD | DOI | main | -0.238 | 0.812 | 0.979 |
|  | BD | DOI | interaction | 0.666 | 0.506 | 0.979 |
|  | SZ | GAF, F | main | 0.699 | 0.485 | 0.979 |
|  | SZ | GAF, F | interaction | 0.234 | 0.815 | 0.979 |
|  | SZ | GAF, S | main | 0.168 | 0.866 | 0.979 |
|  | SZ | GAF, S | interaction | 1.057 | 0.291 | 0.955 |
|  | SZ | score, n | main | -1.297 | 0.195 | 0.843 |
|  | SZ | score, n | interaction | 0.777 | 0.438 | 0.979 |
|  | SZ | score, p | main | 0.608 | 0.543 | 0.979 |
|  | SZ | score, p | interaction | -1.011 | 0.313 | 0.955 |
|  | SZ | AOO | main | -1.777 | 0.076 | 0.617 |
|  | SZ | AOO | interaction | -0.188 | 0.851 | 0.979 |
|  | SZ | DOI | main | -1.086 | 0.278 | 0.955 |
|  | SZ | DOI | interaction | -0.315 | 0.753 | 0.979 |
| Presubiculum | BD | GAF, F | main | 1.010 | 0.313 | 0.955 |
|  | BD | GAF, F | interaction | -0.624 | 0.533 | 0.979 |
|  | BD | GAF, S | main | 0.690 | 0.491 | 0.979 |
|  | BD | GAF, S | interaction | 0.038 | 0.970 | 0.989 |
|  | BD | IDS | main | 0.950 | 0.343 | 0.955 |
|  | BD | IDS | interaction | 0.943 | 0.347 | 0.957 |
|  | BD | YMRS | main | 0.130 | 0.897 | 0.979 |
|  | BD | YMRS | interaction | -0.111 | 0.912 | 0.979 |
|  | BD | AOO | main | -0.752 | 0.453 | 0.979 |
|  | BD | AOO | interaction | -0.955 | 0.340 | 0.955 |
|  | BD | DOI | main | 2.112 | **0.035** | 0.607 |
|  | BD | DOI | interaction | -1.737 | 0.083 | 0.617 |
|  | SZ | GAF, F | main | -1.203 | 0.230 | 0.896 |
|  | SZ | GAF, F | interaction | 0.128 | 0.898 | 0.979 |
|  | SZ | GAF, S | main | -1.036 | 0.301 | 0.955 |
|  | SZ | GAF, S | interaction | 0.281 | 0.779 | 0.979 |
|  | SZ | score, n | main | -0.032 | 0.974 | 0.989 |
|  | SZ | score, n | interaction | -0.362 | 0.718 | 0.979 |
|  | SZ | score, p | main | 2.044 | **0.042** | 0.607 |
|  | SZ | score, p | interaction | -0.361 | 0.718 | 0.979 |
|  | SZ | AOO | main | -1.149 | 0.251 | 0.946 |
|  | SZ | AOO | interaction | -1.361 | 0.174 | 0.828 |
|  | SZ | DOI | main | 0.055 | 0.956 | 0.989 |
|  | SZ | DOI | interaction | 1.557 | 0.120 | 0.767 |
| Subiculum | BD | GAF, F | main | -0.428 | 0.669 | 0.979 |
|  | BD | GAF, F | interaction | -1.848 | 0.066 | 0.617 |
|  | BD | GAF, S | main | -0.378 | 0.706 | 0.979 |
|  | BD | GAF, S | interaction | -0.105 | 0.916 | 0.979 |
|  | BD | IDS | main | 0.501 | 0.616 | 0.979 |
|  | BD | IDS | interaction | 1.741 | 0.083 | 0.617 |
|  | BD | YMRS | main | 0.119 | 0.905 | 0.979 |
|  | BD | YMRS | interaction | -0.554 | 0.580 | 0.979 |
|  | BD | AOO | main | -0.582 | 0.561 | 0.979 |
|  | BD | AOO | interaction | 0.216 | 0.829 | 0.979 |
|  | BD | DOI | main | 2.027 | **0.044** | 0.607 |
|  | BD | DOI | interaction | -2.089 | 0.038 | 0.607 |
|  | SZ | GAF, F | main | -0.811 | 0.418 | 0.979 |
|  | SZ | GAF, F | interaction | 0.578 | 0.563 | 0.979 |
|  | SZ | GAF, S | main | -0.485 | 0.628 | 0.979 |
|  | SZ | GAF, S | interaction | 0.562 | 0.574 | 0.979 |
|  | SZ | score, n | main | -0.180 | 0.857 | 0.979 |
|  | SZ | score, n | interaction | 0.150 | 0.881 | 0.979 |
|  | SZ | score, p | main | 0.894 | 0.372 | 0.979 |
|  | SZ | score, p | interaction | -0.293 | 0.770 | 0.979 |
|  | SZ | AOO | main | 1.979 | **0.048** | 0.607 |
|  | SZ | AOO | interaction | -0.701 | 0.484 | 0.979 |
|  | SZ | DOI | main | 1.518 | 0.130 | 0.774 |
|  | SZ | DOI | interaction | 1.569 | 0.117 | 0.767 |
| Hippocampus | BD | GAF, F | main | 0.953 | 0.341 | 0.955 |
|  | BD | GAF, F | interaction | -0.804 | 0.422 | 0.979 |
|  | BD | GAF, S | main | 0.523 | 0.601 | 0.979 |
|  | BD | GAF, S | interaction | 1.148 | 0.252 | 0.946 |
|  | BD | IDS | main | 0.475 | 0.635 | 0.979 |
|  | BD | IDS | interaction | 0.277 | 0.782 | 0.979 |
|  | BD | YMRS | main | -0.057 | 0.955 | 0.989 |
|  | BD | YMRS | interaction | -1.374 | 0.171 | 0.828 |
|  | BD | AOO | main | -2.570 | **0.011** | 0.332 |
|  | BD | AOO | interaction | -1.051 | 0.294 | 0.955 |
|  | BD | DOI | main | -0.272 | 0.786 | 0.979 |
|  | BD | DOI | interaction | -1.923 | 0.055 | 0.607 |
|  | SZ | GAF, F | main | 0.406 | 0.685 | 0.979 |
|  | SZ | GAF, F | interaction | 0.801 | 0.424 | 0.979 |
|  | SZ | GAF, S | main | 0.904 | 0.367 | 0.979 |
|  | SZ | GAF, S | interaction | 0.303 | 0.762 | 0.979 |
|  | SZ | score, n | main | -0.615 | 0.539 | 0.979 |
|  | SZ | score, n | interaction | -0.538 | 0.591 | 0.979 |
|  | SZ | score, p | main | -0.382 | 0.703 | 0.979 |
|  | SZ | score, p | interaction | -0.574 | 0.566 | 0.979 |
|  | SZ | AOO | main | 0.835 | 0.404 | 0.979 |
|  | SZ | AOO | interaction | -0.834 | 0.405 | 0.979 |
|  | SZ | DOI | main | 0.336 | 0.737 | 0.979 |
|  | SZ | DOI | interaction | 0.499 | 0.618 | 0.979 |

Main effects are based on regression models, adjusted for sex, age, age^2^ and intracranial volume. Interaction effects are based on regression models with a sex-by-clinical measures interaction term, adjusted for age, age^2^ and intracranial volume. As both age at onset (BD r = 0.58, SZ r = 0.66) and duration of illness (BD r = 0.66, SZ r = 0.62) are highly correlated with age, these linear models was only adjusted for intracranial volume. Abbreviation: CA = cornu ammonis, GC-ML-DG = granule cells in the molecular layer of the dentate gyrus, HATA = hippocampal-amygdaloid transition area, GAF = global assessment of function, IDS = inventory for depressive symptomatology, YMRS = young's mania rating scale, FDR = false discover rate. Significant results are highlighted in bold.

**Table S9| Associations between hippocampal volumes and cognitive measures in bipolar disorders (BD) and schizophrenia spectrum disorders (SZ).**

| **Subfields** | **Diagnosis** | **Cognitive domain** | **Effect** | **tvalue** | **pvalue** | **pFDR** |
| --- | --- | --- | --- | --- | --- | --- |
| CA1 | BD | Verbal learning | main | -1.089 | 0.277 | 0.683 |
|  | BD | Verbal learning | interaction | -0.471 | 0.638 | 0.886 |
|  | BD | Verbal memory | main | -0.422 | 0.673 | 0.886 |
|  | BD | Verbal memory | interaction | -0.827 | 0.409 | 0.716 |
|  | BD | Processing speed | main | 1.418 | 0.157 | 0.617 |
|  | BD | Processing speed | interaction | -0.552 | 0.581 | 0.863 |
|  | BD | Working memory | main | 1.284 | 0.200 | 0.619 |
|  | BD | Working memory | interaction | 0.952 | 0.342 | 0.697 |
|  | SZ | Verbal learning | main | -0.005 | 0.996 | 1.000 |
|  | SZ | Verbal learning | interaction | -0.701 | 0.484 | 0.806 |
|  | SZ | Verbal memory | main | -0.228 | 0.820 | 0.942 |
|  | SZ | Verbal memory | interaction | -1.308 | 0.192 | 0.619 |
|  | SZ | Processing speed | main | 1.897 | 0.059 | 0.599 |
|  | SZ | Processing speed | interaction | -1.045 | 0.297 | 0.693 |
|  | SZ | Working memory | main | -0.047 | 0.963 | 0.991 |
|  | SZ | Working memory | interaction | -1.601 | 0.110 | 0.610 |
| CA3 | BD | Verbal learning | main | -0.555 | 0.579 | 0.863 |
|  | BD | Verbal learning | interaction | -1.562 | 0.119 | 0.610 |
|  | BD | Verbal memory | main | -0.116 | 0.908 | 0.983 |
|  | BD | Verbal memory | interaction | -2.060 | **0.040** | 0.524 |
|  | BD | Processing speed | main | 0.425 | 0.671 | 0.886 |
|  | BD | Processing speed | interaction | -0.870 | 0.385 | 0.701 |
|  | BD | Working memory | main | 1.341 | 0.181 | 0.619 |
|  | BD | Working memory | interaction | 1.159 | 0.248 | 0.672 |
|  | SZ | Verbal learning | main | 0.811 | 0.418 | 0.725 |
|  | SZ | Verbal learning | interaction | -0.485 | 0.628 | 0.886 |
|  | SZ | Verbal memory | main | -0.048 | 0.961 | 0.991 |
|  | SZ | Verbal memory | interaction | -1.327 | 0.185 | 0.619 |
|  | SZ | Processing speed | main | -0.117 | 0.907 | 0.983 |
|  | SZ | Processing speed | interaction | -1.239 | 0.216 | 0.633 |
|  | SZ | Working memory | main | 0.000 | 1.000 | 1.000 |
|  | SZ | Working memory | interaction | -0.443 | 0.658 | 0.886 |
| CA4 | BD | Verbal learning | main | -0.560 | 0.576 | 0.863 |
|  | BD | Verbal learning | interaction | -1.081 | 0.281 | 0.683 |
|  | BD | Verbal memory | main | 0.095 | 0.925 | 0.983 |
|  | BD | Verbal memory | interaction | -1.547 | 0.123 | 0.610 |
|  | BD | Processing speed | main | 1.271 | 0.205 | 0.619 |
|  | BD | Processing speed | interaction | -0.632 | 0.528 | 0.838 |
|  | BD | Working memory | main | 0.989 | 0.324 | 0.695 |
|  | BD | Working memory | interaction | 1.471 | 0.143 | 0.617 |
|  | SZ | Verbal learning | main | 1.030 | 0.304 | 0.695 |
|  | SZ | Verbal learning | interaction | -0.383 | 0.702 | 0.896 |
|  | SZ | Verbal memory | main | 0.658 | 0.511 | 0.824 |
|  | SZ | Verbal memory | interaction | -1.522 | 0.129 | 0.610 |
|  | SZ | Processing speed | main | 1.339 | 0.181 | 0.619 |
|  | SZ | Processing speed | interaction | -0.973 | 0.331 | 0.695 |
|  | SZ | Working memory | main | 0.409 | 0.682 | 0.887 |
|  | SZ | Working memory | interaction | -0.694 | 0.488 | 0.806 |
| Fimbria | BD | Verbal learning | main | -1.633 | 0.104 | 0.610 |
|  | BD | Verbal learning | interaction | 0.998 | 0.319 | 0.695 |
|  | BD | Verbal memory | main | -1.504 | 0.134 | 0.610 |
|  | BD | Verbal memory | interaction | 1.352 | 0.177 | 0.619 |
|  | BD | Processing speed | main | 0.187 | 0.852 | 0.967 |
|  | BD | Processing speed | interaction | 1.010 | 0.313 | 0.695 |
|  | BD | Working memory | main | 0.577 | 0.565 | 0.863 |
|  | BD | Working memory | interaction | 2.448 | **0.015** | 0.524 |
|  | SZ | Verbal learning | main | -2.197 | **0.029** | 0.524 |
|  | SZ | Verbal learning | interaction | -1.206 | 0.228 | 0.651 |
|  | SZ | Verbal memory | main | -1.569 | 0.117 | 0.610 |
|  | SZ | Verbal memory | interaction | -1.738 | 0.083 | 0.599 |
|  | SZ | Processing speed | main | 1.544 | 0.123 | 0.610 |
|  | SZ | Processing speed | interaction | 0.871 | 0.384 | 0.701 |
|  | SZ | Working memory | main | -1.350 | 0.178 | 0.619 |
|  | SZ | Working memory | interaction | -1.286 | 0.199 | 0.619 |
| GC-ML-DG | BD | Verbal learning | main | -0.413 | 0.680 | 0.887 |
|  | BD | Verbal learning | interaction | -0.550 | 0.583 | 0.863 |
|  | BD | Verbal memory | main | 0.276 | 0.782 | 0.925 |
|  | BD | Verbal memory | interaction | -1.065 | 0.288 | 0.683 |
|  | BD | Processing speed | main | 1.532 | 0.127 | 0.610 |
|  | BD | Processing speed | interaction | -0.458 | 0.647 | 0.886 |
|  | BD | Working memory | main | 1.420 | 0.157 | 0.617 |
|  | BD | Working memory | interaction | 2.103 | **0.036** | 0.524 |
|  | SZ | Verbal learning | main | 1.097 | 0.273 | 0.683 |
|  | SZ | Verbal learning | interaction | -0.388 | 0.698 | 0.896 |
|  | SZ | Verbal memory | main | 0.852 | 0.395 | 0.707 |
|  | SZ | Verbal memory | interaction | -1.426 | 0.155 | 0.617 |
|  | SZ | Processing speed | main | 1.744 | 0.082 | 0.599 |
|  | SZ | Processing speed | interaction | -0.892 | 0.373 | 0.701 |
|  | SZ | Working memory | main | 0.563 | 0.574 | 0.863 |
|  | SZ | Working memory | interaction | -0.867 | 0.387 | 0.701 |
| HATA | BD | Verbal learning | main | -1.151 | 0.251 | 0.672 |
|  | BD | Verbal learning | interaction | 0.181 | 0.857 | 0.967 |
|  | BD | Verbal memory | main | -0.865 | 0.388 | 0.701 |
|  | BD | Verbal memory | interaction | -0.466 | 0.642 | 0.886 |
|  | BD | Processing speed | main | 0.263 | 0.792 | 0.926 |
|  | BD | Processing speed | interaction | -0.671 | 0.503 | 0.817 |
|  | BD | Working memory | main | 2.257 | **0.025** | 0.524 |
|  | BD | Working memory | interaction | 1.569 | 0.118 | 0.610 |
|  | SZ | Verbal learning | main | -1.270 | 0.205 | 0.619 |
|  | SZ | Verbal learning | interaction | -1.637 | 0.102 | 0.610 |
|  | SZ | Verbal memory | main | -1.171 | 0.242 | 0.672 |
|  | SZ | Verbal memory | interaction | -1.786 | 0.075 | 0.599 |
|  | SZ | Processing speed | main | 0.546 | 0.585 | 0.863 |
|  | SZ | Processing speed | interaction | -1.131 | 0.259 | 0.682 |
|  | SZ | Working memory | main | 0.743 | 0.458 | 0.781 |
|  | SZ | Working memory | interaction | -1.147 | 0.252 | 0.672 |
| Fissure | BD | Verbal learning | main | 0.474 | 0.636 | 0.886 |
|  | BD | Verbal learning | interaction | -2.060 | **0.040** | 0.524 |
|  | BD | Verbal memory | main | 0.798 | 0.425 | 0.731 |
|  | BD | Verbal memory | interaction | -2.109 | **0.036** | 0.524 |
|  | BD | Processing speed | main | -0.170 | 0.865 | 0.968 |
|  | BD | Processing speed | interaction | -0.985 | 0.326 | 0.695 |
|  | BD | Working memory | main | 1.737 | 0.084 | 0.599 |
|  | BD | Working memory | interaction | 0.143 | 0.886 | 0.981 |
|  | SZ | Verbal learning | main | -2.125 | **0.034** | 0.524 |
|  | SZ | Verbal learning | interaction | -1.228 | 0.220 | 0.636 |
|  | SZ | Verbal memory | main | -2.583 | **0.010** | 0.524 |
|  | SZ | Verbal memory | interaction | -0.336 | 0.737 | 0.918 |
|  | SZ | Processing speed | main | -1.850 | 0.065 | 0.599 |
|  | SZ | Processing speed | interaction | -1.937 | 0.053 | 0.586 |
|  | SZ | Working memory | main | -1.251 | 0.212 | 0.629 |
|  | SZ | Working memory | interaction | -0.616 | 0.538 | 0.848 |
| Tail | BD | Verbal learning | main | -0.149 | 0.882 | 0.981 |
|  | BD | Verbal learning | interaction | -0.440 | 0.660 | 0.886 |
|  | BD | Verbal memory | main | 0.897 | 0.371 | 0.701 |
|  | BD | Verbal memory | interaction | -0.955 | 0.341 | 0.697 |
|  | BD | Processing speed | main | 1.330 | 0.185 | 0.619 |
|  | BD | Processing speed | interaction | 1.287 | 0.199 | 0.619 |
|  | BD | Working memory | main | 0.111 | 0.912 | 0.983 |
|  | BD | Working memory | interaction | 1.063 | 0.289 | 0.683 |
|  | SZ | Verbal learning | main | 1.108 | 0.268 | 0.683 |
|  | SZ | Verbal learning | interaction | -0.866 | 0.387 | 0.701 |
|  | SZ | Verbal memory | main | 0.321 | 0.748 | 0.921 |
|  | SZ | Verbal memory | interaction | -2.136 | **0.033** | 0.524 |
|  | SZ | Processing speed | main | 0.531 | 0.596 | 0.873 |
|  | SZ | Processing speed | interaction | -1.269 | 0.205 | 0.619 |
|  | SZ | Working memory | main | 0.093 | 0.926 | 0.983 |
|  | SZ | Working memory | interaction | -1.956 | 0.051 | 0.586 |
| Molecular layer | BD | Verbal learning | main | -0.893 | 0.373 | 0.701 |
|  | BD | Verbal learning | interaction | -0.468 | 0.640 | 0.886 |
|  | BD | Verbal memory | main | -0.077 | 0.939 | 0.986 |
|  | BD | Verbal memory | interaction | -0.891 | 0.373 | 0.701 |
|  | BD | Processing speed | main | 1.497 | 0.135 | 0.610 |
|  | BD | Processing speed | interaction | -0.567 | 0.571 | 0.863 |
|  | BD | Working memory | main | 0.890 | 0.374 | 0.701 |
|  | BD | Working memory | interaction | 0.979 | 0.328 | 0.695 |
|  | SZ | Verbal learning | main | 0.703 | 0.483 | 0.806 |
|  | SZ | Verbal learning | interaction | -0.356 | 0.722 | 0.910 |
|  | SZ | Verbal memory | main | 0.311 | 0.756 | 0.921 |
|  | SZ | Verbal memory | interaction | -0.963 | 0.336 | 0.697 |
|  | SZ | Processing speed | main | 2.185 | **0.029** | 0.524 |
|  | SZ | Processing speed | interaction | -1.170 | 0.243 | 0.672 |
|  | SZ | Working memory | main | 0.463 | 0.644 | 0.886 |
|  | SZ | Working memory | interaction | -1.076 | 0.283 | 0.683 |
| Parasubiculum | BD | Verbal learning | main | -0.252 | 0.801 | 0.927 |
|  | BD | Verbal learning | interaction | 1.780 | 0.076 | 0.599 |
|  | BD | Verbal memory | main | -0.304 | 0.762 | 0.921 |
|  | BD | Verbal memory | interaction | 1.075 | 0.283 | 0.683 |
|  | BD | Processing speed | main | 0.451 | 0.653 | 0.886 |
|  | BD | Processing speed | interaction | -0.102 | 0.919 | 0.983 |
|  | BD | Working memory | main | 1.310 | 0.191 | 0.619 |
|  | BD | Working memory | interaction | 0.400 | 0.689 | 0.891 |
|  | SZ | Verbal learning | main | 1.791 | 0.074 | 0.599 |
|  | SZ | Verbal learning | interaction | 0.604 | 0.546 | 0.854 |
|  | SZ | Verbal memory | main | 1.791 | 0.074 | 0.599 |
|  | SZ | Verbal memory | interaction | 0.294 | 0.769 | 0.925 |
|  | SZ | Processing speed | main | 2.274 | **0.023** | 0.524 |
|  | SZ | Processing speed | interaction | 0.309 | 0.758 | 0.921 |
|  | SZ | Working memory | main | 3.169 | **0.002** | 0.344 |
|  | SZ | Working memory | interaction | 0.825 | 0.410 | 0.716 |
| Presubiculum | BD | Verbal learning | main | -0.128 | 0.898 | 0.983 |
|  | BD | Verbal learning | interaction | 1.093 | 0.275 | 0.683 |
|  | BD | Verbal memory | main | 0.278 | 0.781 | 0.925 |
|  | BD | Verbal memory | interaction | 0.429 | 0.668 | 0.886 |
|  | BD | Processing speed | main | 1.498 | 0.135 | 0.610 |
|  | BD | Processing speed | interaction | 0.047 | 0.963 | 0.991 |
|  | BD | Working memory | main | 0.454 | 0.650 | 0.886 |
|  | BD | Working memory | interaction | 0.997 | 0.319 | 0.695 |
|  | SZ | Verbal learning | main | 1.448 | 0.148 | 0.617 |
|  | SZ | Verbal learning | interaction | -0.093 | 0.926 | 0.983 |
|  | SZ | Verbal memory | main | 1.696 | 0.091 | 0.610 |
|  | SZ | Verbal memory | interaction | 0.056 | 0.956 | 0.991 |
|  | SZ | Processing speed | main | 2.574 | **0.010** | 0.524 |
|  | SZ | Processing speed | interaction | -0.250 | 0.803 | 0.927 |
|  | SZ | Working memory | main | 2.319 | **0.021** | 0.524 |
|  | SZ | Working memory | interaction | -0.325 | 0.745 | 0.921 |
| Subiculum | BD | Verbal learning | main | -0.893 | 0.372 | 0.701 |
|  | BD | Verbal learning | interaction | 0.426 | 0.670 | 0.886 |
|  | BD | Verbal memory | main | -0.014 | 0.989 | 1.000 |
|  | BD | Verbal memory | interaction | -0.367 | 0.714 | 0.906 |
|  | BD | Processing speed | main | 0.638 | 0.524 | 0.838 |
|  | BD | Processing speed | interaction | -0.210 | 0.834 | 0.953 |
|  | BD | Working memory | main | -0.176 | 0.860 | 0.967 |
|  | BD | Working memory | interaction | 0.675 | 0.500 | 0.817 |
|  | SZ | Verbal learning | main | -0.278 | 0.781 | 0.925 |
|  | SZ | Verbal learning | interaction | -1.029 | 0.304 | 0.695 |
|  | SZ | Verbal memory | main | -0.038 | 0.970 | 0.994 |
|  | SZ | Verbal memory | interaction | -1.333 | 0.183 | 0.619 |
|  | SZ | Processing speed | main | 2.090 | **0.037** | 0.524 |
|  | SZ | Processing speed | interaction | -1.530 | 0.127 | 0.610 |
|  | SZ | Working memory | main | -0.005 | 0.996 | 1.000 |
|  | SZ | Working memory | interaction | -1.425 | 0.155 | 0.617 |
| Hippocampus | BD | Verbal learning | main | -0.889 | 0.375 | 0.701 |
|  | BD | Verbal learning | interaction | -0.267 | 0.790 | 0.926 |
|  | BD | Verbal memory | main | 0.022 | 0.983 | 1.000 |
|  | BD | Verbal memory | interaction | -0.895 | 0.371 | 0.701 |
|  | BD | Processing speed | main | 1.503 | 0.134 | 0.610 |
|  | BD | Processing speed | interaction | -0.085 | 0.932 | 0.984 |
|  | BD | Working memory | main | 0.981 | 0.328 | 0.695 |
|  | BP | Working memory | interaction | 1.489 | 0.138 | 0.610 |
|  | SZ | Verbal learning | main | 0.696 | 0.487 | 0.806 |
|  | SZ | Verbal learning | interaction | -0.838 | 0.402 | 0.715 |
|  | SZ | Verbal memory | main | 0.341 | 0.733 | 0.918 |
|  | SZ | Verbal memory | interaction | -1.782 | 0.076 | 0.599 |
|  | SZ | Processing speed | main | 2.028 | **0.043** | 0.528 |
|  | SZ | Processing speed | interaction | -1.322 | 0.187 | 0.619 |
|  | SZ | Working memory | main | 0.500 | 0.617 | 0.886 |
|  | SZ | Working memory | interaction | -1.602 | 0.110 | 0.610 |

Main effects are based on regression models, adjusted for sex, age, age^2^ and intracranial volume. Interaction effects are based on regression models with a sex-by-clinical measures interaction term, adjusted for age, age^2^ and intracranial volume. Abbreviation: CA = cornu ammonis, GC-ML-DG = granule cells in the molecular layer of the dentate gyrus, HATA = hippocampal-amygdaloid transition area, FDR = false discover rate. Significant results are highlighted in bold.
